# Mitigation of SARS-CoV-2 Transmission at a Large Public University

**DOI:** 10.1101/2021.08.03.21261548

**Authors:** Diana Rose E. Ranoa, Robin L. Holland, Fadi G. Alnaji, Kelsie J. Green, Leyi Wang, Richard L. Fredrickson, Tong Wang, George N. Wong, Johnny Uelmen, Sergei Maslov, Ahmed Elbanna, Zachary J. Weiner, Alexei V. Tkachenko, Hantao Zhang, Zhiru Liu, Sanjay J. Patel, John M. Paul, Nickolas P. Vance, Joseph G. Gulick, Sandeep Puthanveetil Satheesan, Isaac J. Galvan, Andrew Miller, Joseph Grohens, Todd J. Nelson, Mary P. Stevens, P. Mark Hennessy, Robert C. Parker, Edward Santos, Charles Brackett, Julie D. Steinman, Melvin R. Fenner, Kirstin Dohrer, Kraig Wagenecht, Michael DeLorenzo, Laura Wilhelm-Barr, Brian R. Brauer, Catherine Best-Popescu, Gary Durack, Nathan Wetter, David M. Kranz, Jessica Breitbarth, Charlie Simpson, Julie A. Pryde, Robin N. Kaler, Chris Harris, Allison C. Vance, Jodi L. Silotto, Mark Johnson, Enrique Valera, Patricia K. Anton, Lowa Mwilambwe, Stephen P. Bryan, Deborah S. Stone, Danita B. Young, Wanda E. Ward, John Lantz, John A. Vozenilek, Rashid Bashir, Jeffrey S. Moore, Mayank Garg, Julian C. Cooper, Gillian Snyder, Michelle H. Lore, Dustin L. Yocum, Neal J. Cohen, Jan E. Novakofski, Melanie J. Loots, Randy L. Ballard, Mark Band, Kayla M. Banks, Joseph D. Barnes, Iuliana Bentea, Jessica Black, Jeremy Busch, Hannah Christensen, Abigail Conte, Madison Conte, Michael Curry, Jennifer Eardley, April Edwards, Therese Eggett, Judes Fleurimont, Delaney Foster, Bruce W. Fouke, Nicholas Gallagher, Nicole Gastala, Scott A. Genung, Declan Glueck, Brittani Gray, Andrew Greta, Robert M. Healy, Ashley Hetrick, Arianna A Holterman, Nahed Ismail, Ian Jasenof, Patrick Kelly, Aaron Kielbasa, Teresa Kiesel, Lorenzo M. Kindle, Rhonda L. Lipking, Yukari C. Manabe, Jade Mayes, Reubin McGuffin, Kenton G. McHenry, Agha Mirza, Jada Moseley, Heba H. Mostafa, Melody Mumford, Kathleen Munoz, Arika D. Murray, Moira Nolan, Nil A. Parikh, Andrew Pekosz, Janna Pflugmacher, Janise M. Phillips, Collin Pitts, Mark C. Potter, James Quisenberry, Janelle Rear, Matthew L. Robinson, Edith Rosillo, Leslie N. Rye, MaryEllen Sherwood, Anna Simon, Jamie M. Singson, Carly Skadden, Tina H. Skelton, Charlie Smith, Mary Stech, Ryan Thomas, Matthew A. Tomaszewski, Erika A. Tyburski, Scott Vanwingerden, Evette Vlach, Ronald S. Watkins, Karriem Watson, Karen C. White, Timothy L. Killeen, Robert J. Jones, Andreas C. Cangellaris, Susan A. Martinis, Awais Vaid, Christopher B. Brooke, Joseph T. Walsh, William C. Sullivan, Rebecca L. Smith, Nigel D. Goldenfeld, Timothy M. Fan, Paul J. Hergenrother, Martin D. Burke

## Abstract

In the Fall of 2020, many universities saw extensive transmission of SARS-CoV-2 among their populations, threatening the health of students, faculty and staff, the viability of in-person instruction, and the health of surrounding communities.^1, 2^ Here we report that a multimodal “SHIELD: Target, Test, and Tell” program mitigated the spread of SARS-CoV-2 at a large public university, prevented community transmission, and allowed continuation of in-person classes amidst the pandemic. The program combines epidemiological modelling and surveillance (Target); fast and frequent testing using a novel and FDA Emergency Use Authorized low-cost and scalable saliva-based RT-qPCR assay for SARS-CoV-2 that bypasses RNA extraction, called covidSHIELD (Test); and digital tools that communicate test results, notify of potential exposures, and promote compliance with public health mandates (Tell). These elements were combined with masks, social distancing, and robust education efforts. In Fall 2020, we performed more than 1,000,000 covidSHIELD tests while keeping classrooms, laboratories, and many other university activities open. Generally, our case positivity rates remained less than 0.5%, we prevented transmission from our students to our faculty and staff, and data indicate that we had no spread in our classrooms or research laboratories. During this fall semester, we had zero COVID-19-related hospitalizations or deaths amongst our university community. We also prevented transmission from our university community to the surrounding Champaign County community. Our experience demonstrates that multimodal transmission mitigation programs can enable university communities to achieve such outcomes until widespread vaccination against COVID-19 is achieved, and provides a roadmap for how future pandemics can be addressed.

---

As our ∼35,000 undergraduate students at the University of Illinois at Urbana-Champaign (UIUC) departed campus to continue their education remotely in Spring of 2020, we recognized their return in the Fall would present significant challenges. Our biggest concern was that unmitigated transmission amongst our undergraduate student population would drive increased cases in our faculty and staff and/or the surrounding Champaign County community. The SHIELD program was devised to mitigate SARS-CoV-2 transmission through identification and safe isolation of infected individuals before they spread the virus. The Target, Test, and Tell components of SHIELD are used synergistically and allowed us to achieve our transmission mitigation goal.

## Target

For the *Target* component of the SHIELD program, epidemiological modelling helped determine who should be tested and how often, and real-time data analysis further allowed for strategic adjustments to testing schedules throughout the semester to maximally mitigate spread. Given the data suggesting that SARS-CoV-2 can be transmitted by pre-symptomatic and asymptomatic carriers,^3–8^ fast and frequent testing of all individuals in the university community was expected to be critical for mitigating localized outbreaks by enabling identification and isolation of infected individuals prior to clinically-impactful shedding of SARS-CoV-2.^9, 10^ Available data on viral dynamics^8, 9, 11, 12^ suggested that test results should be returned within hours, not days, and that testing might need to be performed multiple times per week, particularly for the populations most likely to be exposed to SARS-CoV-2. To explore these issues quantitatively, we used a variety of methods to arrive at an optimal strategy for our campus.

We calculated how the basic reproduction number, R_o_, is modified by a multiplier, *M*, that accounts for the fact that if an individual is detected to be positive and immediately isolated, they are unable to continue infecting others. This results in a fractional reduction of R_o_ (R_t_ = *M* R_o_) as detailed in Fig. 1a. Using an infectivity profile that includes pre-symptomatic shedding,^8, 13^ we could calculate *M* as a function of testing frequency (see Methods). We found that testing everyone every 7 days yields *M = 0.71*, but testing everyone every 3.5 days yields *M = 0.45*, because their infectious period while not isolated (Area A in Fig. 1a) is reduced.

**Figure 1.**
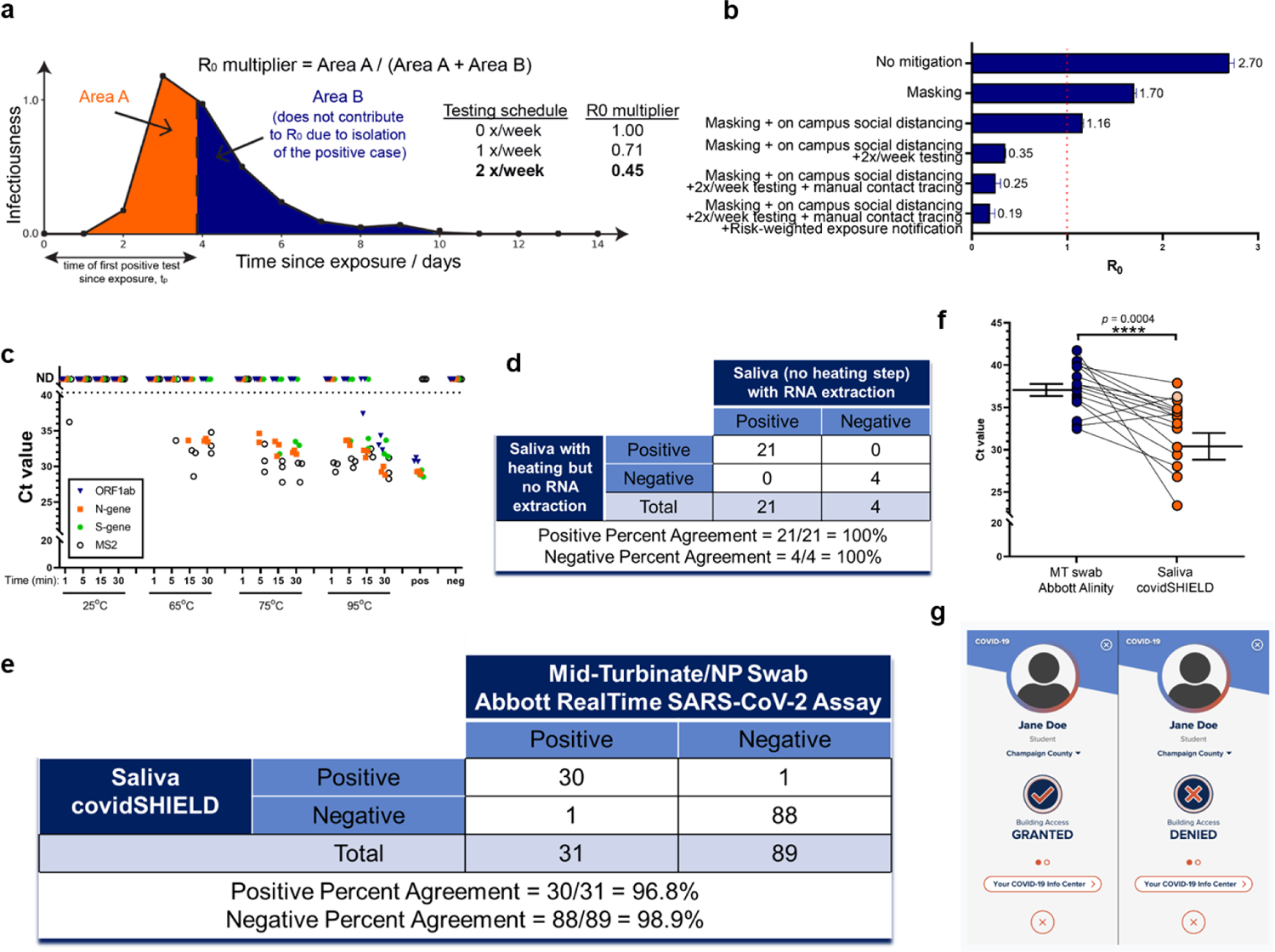
Target, Test, and Tell. **a**, Sensitive testing can reveal a positive case early in the infection, and thus isolation of the index case reduces the number of people infected by this index case. Frequent testing and rapid isolation reduce the time period during which a person is infectious but not isolated (Area A). As a result, the R_0_ multiplier for testing is the ratio between the truncated area under the curve (Area A) and the untruncated area under the curve (Area A + Area B). The dashed vertical line between Area A and Area B represents the moment an infected individual is isolated; as this line moves to the left, *M* is decreased and viral spread is reduced. **b,** Effect of different mitigation interventions on the basic reproduction number R_0_ as computed in our agent-based model. If R_0_ is greater than one (orange dashed line at R_0_=1), the epidemic grows exponentially. If R_0_ is less than one, any outbreak diminishes exponentially. Without any mitigation, R_0_ is close to 3 and a runaway epidemic will occur. Masking and social distancing help reduce transmission but can’t suppress growth of cases on their own as R_0_ is still greater than one. However, when these measures are combined with frequent testing (2 tests a week), R_0_ drops to 0.35 and containment of epidemic becomes possible. Adding extra mitigation interventions such as manual contact tracing and risk based exposure notification being R_0_ further down to 0.19 suggesting the potential for strong control of the epidemic on campus. The details of the agent-based model are given in Supplementary Materials. The results shown here are computed assuming that 100% of the students are compliant with twice a week testing, isolation, and quarantine. We also ran the same simulation assuming 60% compliance, and the same general trends were observed with R_0_ for the full SHIELD program predicted to still be manageable (0.5, see Extended Data Fig. 1). **c,** The effect of heat on SARS-CoV-2 nucleic acid detection in saliva. γ-irradiated SARS-CoV-2 (from BEI, used at 1.0×10^4^ viral copies/mL) was spiked into fresh human saliva (SARS-CoV-2 negative). Samples diluted 1:1 with 2X Tris-borate-EDTA (TBE) buffer (0.5 mL in 50 mL conical tubes) were incubated at 25°C (ambient temperature), or in a hot water bath at 65°C, 75°C, or 95°C, for 1, 5, 15, or 30 min. All saliva samples were spiked with purified MS2 bacteriophage (1:40 MS2:sample) as an internal control. Virus-spiked saliva samples, a positive control (pos; SARS-CoV-2 positive control, 5.0×10^3^ copies/mL, no MS2) and a negative control (neg; water, no MS2) were directly analyzed by RT-qPCR, in triplicate, for SARS-CoV-2 nucleic acid corresponding with ORF1ab gene (blue triangle), N-gene (orange square), and S-gene (green circle), and MS2 (open circle). Undetermined Ct values are plotted as ND. This experiment was repeated at least three times. **d**, 25 clinical saliva samples were split into two aliquots upon receipt, one set was processed using our covidSHIELD assay and the other set was subjected to RNA extraction using MagMax Viral/Pathogen II (MVP II) Nucleic Acid Isolation Kit (ThermoFisher). 5uL of either processed saliva (in 1:1 2x TBE/Tween-20 buffer) or purified RNA (in water) were subsequently used as templates for RT-qPCR. A positive result is called when two out of three viral target genes is detected. **e,** Qualitative outcome of parallel testing of paired mid-turbinate swabs and saliva with the Abbott RealTime SARS-CoV-2 assay and covidSHIELD. A total of 120 participants were enrolled in a clinical study comparing results from contemporaneously collected nasopharyngeal or mid-turbinate nasal swabs analyzed by an FDA Emergency Use Authorized reference method for detection of SARS-CoV-2 and saliva samples analyzed by covidSHIELD. Overall concordance was 98.3% (95% CI, 94.1-99.8%), positive percent agreement was 96.8% (95% CI, 83.2-99.9%) and negative percent agreement was 98.9% (95% CI, 93.9-99.9%). All clinical trials were reviewed by the Western Institutional Review Board. All participants gave written and informed consent. **f,** Additional clinical study outcome of 17 individuals confirmed to be positive for COVID-19 and to have low viral loads (Ct = 32-42, average 37) by mid-turbinate nasal swabs analyzed at the Johns Hopkins University School of Medicine using Abbott Alinity compared with contemporaneously collected saliva samples that were analyzed using the covidSHIELD assay at the University of Illinois Urbana-Champaign CLIA-registered laboratory. *p*-value=0.0004 was calculated using 2-tailed, unpaired t-test. **g,** Mock representative images from the Safer Illinois app demonstrate how compliance with the SHIELD testing protocol was coupled to building access. The screen on the left appears when a user is in compliance with the campus testing protocol and has received a recent negative test for SARS-CoV-2. The screen on the right appears when the user of the app is out of compliance, when they have had a recent exposure notification, or when they have tested positive for the virus. The area within the upper circle containing silhouette image typically displays a picture of the user of the app, one of a number of security features built into Safer Illinois.

We also simulated the spread of COVID-19 on campus using agent-based modelling, following each student as they attend university activities and participate in off-campus socializing (see Methods for key assumptions and details). As shown in Fig. 1b, this analysis predicted that masking or the combination of masking and social distancing would provide inadequate protection against spreading of COVID-19 (R_0_ > 1). However, the addition of twice per week testing was predicted to make R_0_ manageable (R_0_ < 1). The additional inclusion of testing-enabled manual contact tracing and digital exposure notifications predicted even further reductions. In fact, the model predicted these mitigation approaches would be synergistic and highly effective when applied in concert, with R_0_ small enough to contain the epidemic when using the full suite of twice-weekly testing of everyone on campus, isolation of newly infected people, contact tracing, quarantining and use of the Safer Illinois exposure-notification app, along with masking and social distancing. These general trends were robust within the validated boundary conditions of the underlying assumptions (see Methods and Extended Data Fig. 1). These simulations reinforced that the effectiveness of testing, isolation and contact tracing in reducing transmission was heavily dependent on rapid turnaround, because of the high transmissibility of COVID-19, in agreement with other modeling results^14^.

## Test

When considering various testing options, the evaluation of virus levels in saliva was highly attractive due to the known detection of SARS-CoV-2 through oral shedding and the potential for rapid, easy, and non-invasive self-collection,^15–17^ thus minimizing the need for direct healthcare provider-patient contact and consequent conservation of personal protective equipment (PPE). Numerous reports have detailed the detection of SARS-CoV-2 in saliva,^18–22^ and salivary/respiratory aerosols and droplets are recognized as a significant factor in person-to-person transmission of SARS-CoV-2.^15^ However, all saliva-based assays available in the Spring of 2020 required RNA isolation, which added cost, time, and supply chain bottlenecks.

With the goal of performing up to 20,000 individual RT-qPCR tests for SARS-CoV-2 per day, we developed a saliva-based assay for SARS-CoV-2 nucleic acid detection that bypasses RNA isolation/purification^23^ (called covidSHIELD). This process relies on up-front heating of freshly collected saliva samples, an attractive and simple method to inactivate the virus without having to open the collection vessel. Using intact, γ-irradiated SARS-CoV-2 spiked into fresh human saliva (that was confirmed to be SARS-CoV-2 negative), we observed substantial time- and temperature-dependent improvement in SARS-CoV-2 nucleic acid detection by direct RT-qPCR, without the use of RNA extraction (Fig. 1c). When incubated at ambient temperature (no heat treatment), no SARS-CoV-2 genes are detectable, but as temperature and incubation time increase, substantial improvement in viral nucleic acid detection is observed, with 100% identification of the three targeted SARS-CoV-2 genes, in all replicate samples, detected following a 30 minute incubation at 95°C. Importantly, a short heating time (5 minutes) at 95°C (as has been examined by others^24, 25^) does not allow for sensitive detection; the 30-minute duration is essential, as it is likely that this extended heating drives temperature dependent inactivation of components of saliva that inhibit RT-qPCR. Preliminary comparison of this heating-based RNA extraction-free protocol to a standard protocol involving RNA isolation using 25 split clinical saliva samples showed 100% concordance between the two assays (Fig. 1d and Extended Data Table 1), suggesting that the heat-inactivation step does not affect assay outcome. This RNA extraction-free protocol, which is based upon the detection of three viral genes (ORF1ab, N-gene, and S-gene) using the TaqPath COVID-19 Combo kit is also highly sensitive (Limit of Detection of 500-1000 SARS-CoV-2 viral copies/mL, Extended Data Fig. 2), can be optimized for high-throughput using robotic sample transfers with no impact on sensitivity (Extended Data Table 2), is minimally affected by exogenous and endogenous potentially interfering substances (Extended Data Table 3), yields stable results for up to 7 days when the saliva is stored below 25°C prior to heat inactivation (Extended Data Figure 3), and is also compatible with other RT-qPCR primers (such as the N1/N2 CDC primers, Extended Data Fig. 4). Head-to-head comparison with a subsequently developed direct saliva to RT-qPCR technique that requires opening of tubes and addition of proteinase-K to saliva samples prior to heat inactivation^26^ shows that the simple covidSHIELD protocol for viral inactivation results in approximately 8-fold more sensitive detection using the TaqPath COVID-19 Combo kit (Extended Data Fig. 5).

**Figure 2.**
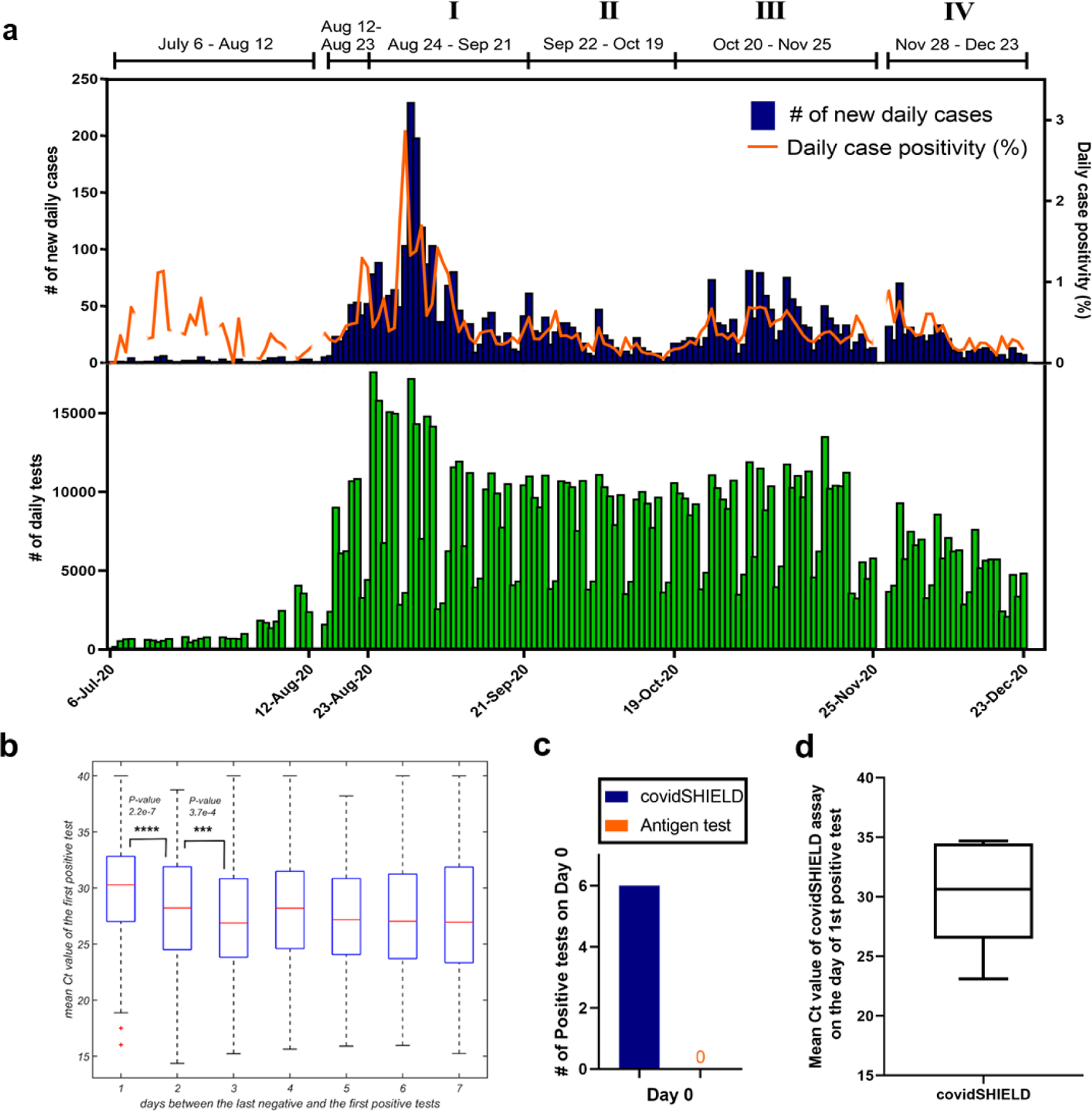
Deployment of frequent repeat testing at the University of Illinois in fall 2020. **a**, Timeline of detected cases during surveillance testing from July 6 to December 23, 2020.The top panel displays the daily new cases (blue) and the daily case positivity (orange). The daily case positivity is computed as the (number of new cases)/(unique number of individuals tested during the day). The lower panel shows the number of daily tests performed (green), which to an excellent approximation is the same as the number of unique individuals tested in a day. **b,** Ct values of the first positive test as a function of time elapsed since the last negative test. The difference between the first (1 day) and the second (2 days) bins is highly statistically significant (*p*-value 2.2e-7), and that between the second (2 days) and the third (3 days) bins is statistically significant (*p*-value 3.7e-4). *P*-values shown are for the two-sided hypothesis of non-zero Pearson correlation between the number of days since the last negative test (x-axis) and the Ct value of the first positive test (y-axis). The exact sample sizes are: 341 patients who test positive 1 day since the last negative, 616 patients - 2 days since the last negative, 716 patients - 3 days since the last negative, and 1230 patients - between 4 and 7 days since the last negative. **c,** Head-to-head daily testing with covidSHIELD and antigen-based lateral flow assays in a subgroup of participants (n=190) from October 1, 2020 to Dec 23, 2020. A total of 13,299 contemporaneous tests were performed. Of the 190 individuals, 6 tested positive for SARS-CoV-2 on Day 0 using the covidSHIELD test but all six tested negative using the antigen test. Blue and orange bars represent the percentage of participants that tested positive for SARS-CoV-2 on day 0 using covidSHIELD and the antigen test, respectively. **d**, Mean Ct vlues of the 6 individuals who tested positive for SARS-CoV-2 using the covidSHIELD assay on Day 0.

**Figure 3.**
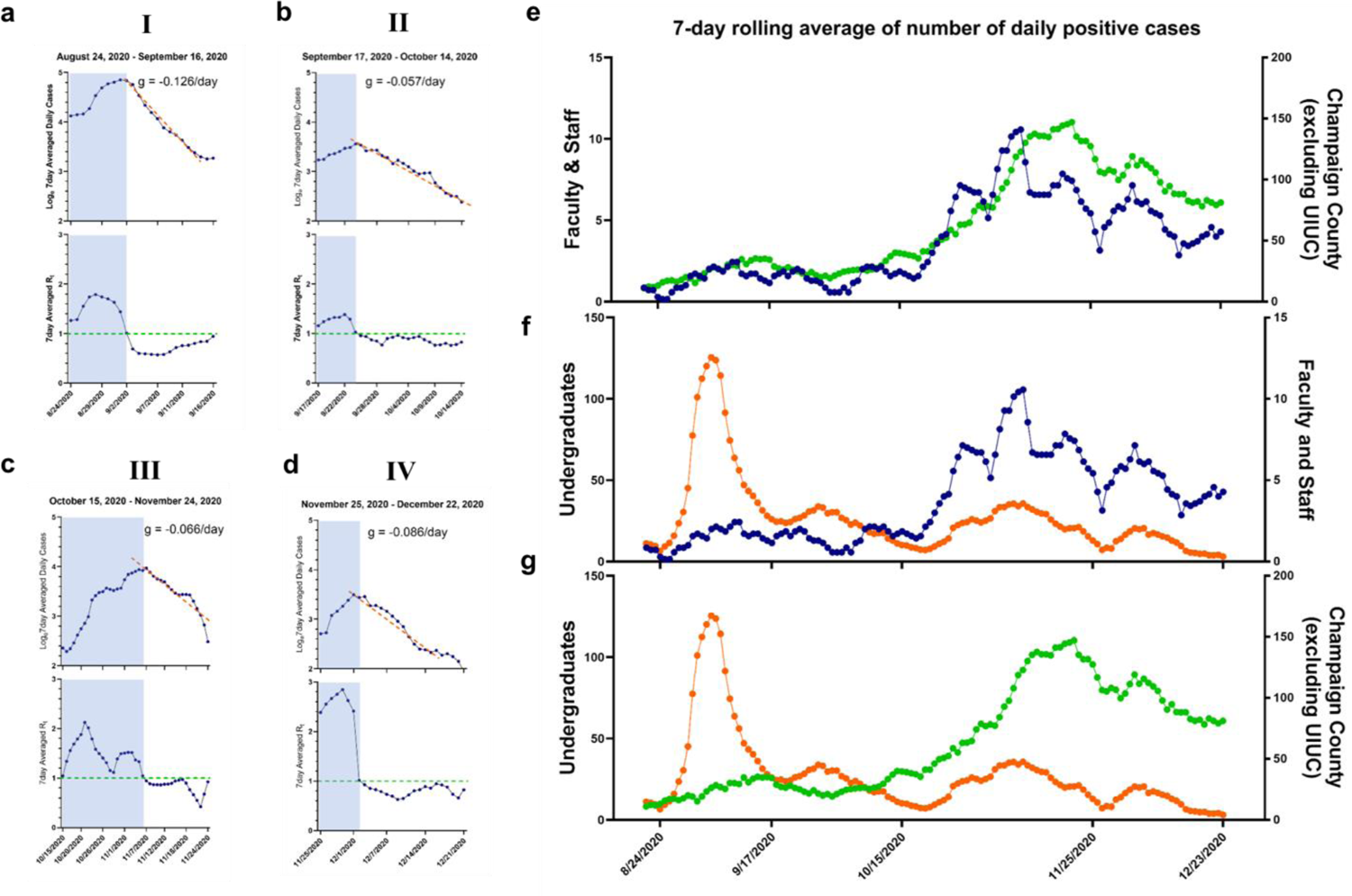
Mitigation of SARS-CoV-2 spread. **a-d**, Effective reproduction number R_t_ during the four periods of time during which transient increases in COVID-19 cases were observed. In each dataset, the top panel displays log_e_ of 7-day averaged daily cases. The dashed lines represent fits for the epidemic curve when the cases were decreasing exponentially and the magnitude of the exponential decay is highlighted on the figure during the different time periods. The decay rates were used to estimate the effective reproduction number using two methods: Wallinga and Lipsitch^32^ that assumes a gamma distribution for the generation interval, and a simple SIR model that assumes an exponential distribution. The estimated values for the different periods shown above are: a. (0.5465, 0.874); b. (0.7837, 0.943);c. (0.751, 0.934); and d. (0.681, 0.914) where the first number in the brackets corresponds to the method of Wallinga and Lipchitz and the second number corresponds to an SIR model. The bottom panel shows the 7-day averaged effective reproduction number Rt computed using the daily new cases according to the method of Cori et al.^33^ The estimates for the reproduction number from the different methods are thus broadly consistent. The shaded regions correspond to dates during which the effective reproduction number rose transiently above 1. **e-g,** The daily number of 7-day averaged daily new cases between faculty/staff and residents in Champaign County **e**, undergraduates and faculty/staff **f**, and undergraduate students and residents in Champaign County **g**, for the period between August 15 and December 23. All points in three plots are colored according to their categories (orange: undergraduate students, blue: faculty/staff, green: residents in Champaign County). Pearson correlation coefficient, 95% confidence interval, and *p*-values for two-tailed test were calculated using GraphPad Prism software.

**Figure 4.**
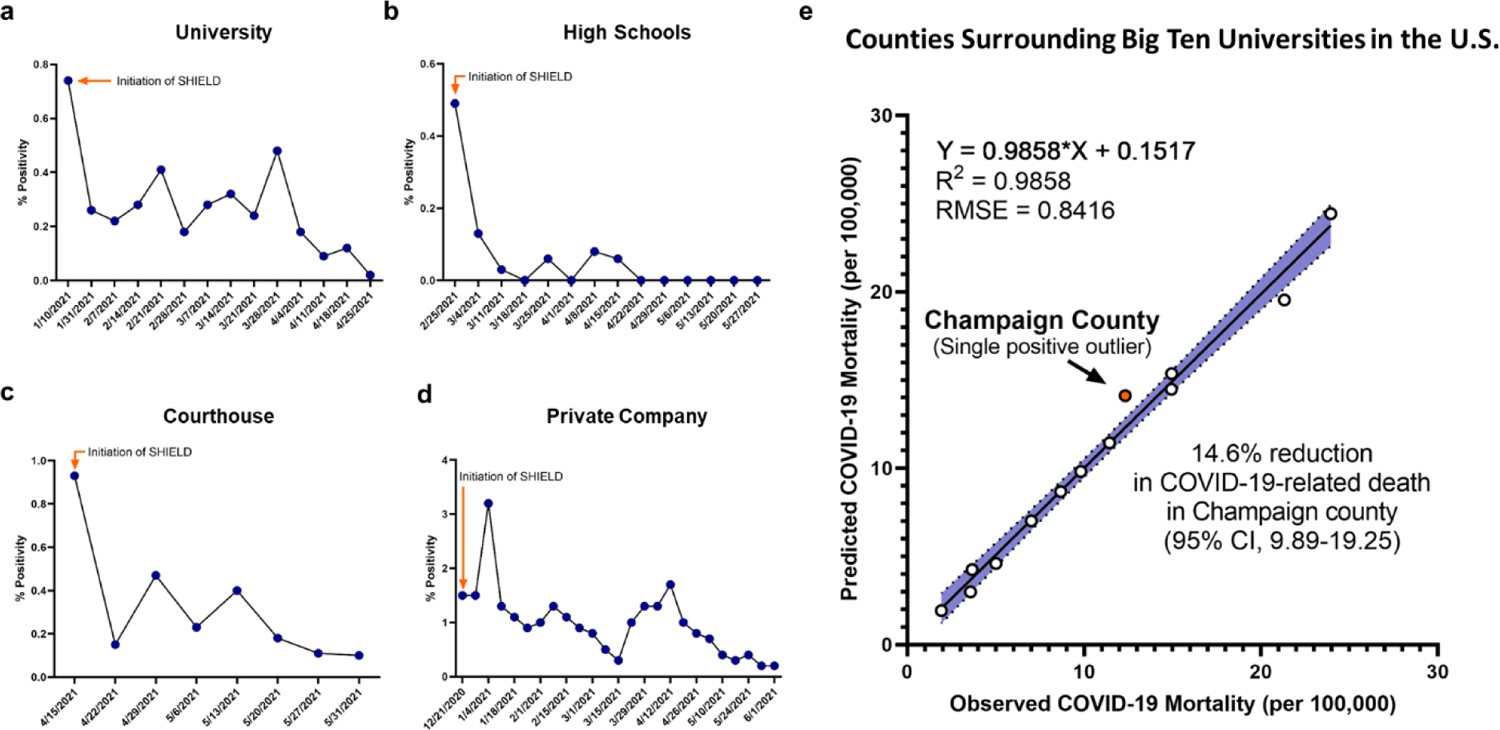
Other communities with and without SHIELD: SHIELD was deployed at other campuses in Winter 2020/Spring 2021. The results from four representative examples are shown: **a**, a university (1x/week), **b,** a pair of high schools (2x/week), **c,** a courthouse (2x/week), and **d**, a large private corporate campus (2x/week). **e,** Relationship between observed and predicted COVID-19 mortality among Big 10 University counties (n=13*). Predicted COVID-19 mortality was analyzed using each county’s social vulnerability index (SVI) and race- and age-adjusted COVID-19 mortality, accounting for state (due to policy differences in COVID-19 management). COVID-19 human case data^34^ and SVI were provided from CDC (2021) and population data was provided from the U.S. Census Bureau (2019).^35^ The line indicates the fit line, with shaded area indicating the 95% confidence intervals around the fit and circles indicating observations with Champaign County shown in orange. *Hennepin and Ramsey Counties (MN) were combined for University of Minnesota; 2 Big 10 University counties (Lancaster, NE and Prince George’s County, MD) did not report COVID-19 mortality for 2020, and were excluded.

In parallel to the work on test development, we created a dedicated CLIA-registered laboratory in our on-campus Veterinary Diagnostic Laboratory that includes semi-robotic capacity to efficiently process up to 20,000 saliva samples per day. We also set up ∼20 saliva collection stations throughout campus and all associated infrastructure for hourly delivery of samples to the CLIA lab.

A clinical study was performed with 120 individuals suspected of COVID-19 by their healthcare provider to compare the results of the covidSHIELD assay to results from contemporaneously collected nasopharyngeal or mid-turbinate nasal swabs analyzed by an FDA Emergency Use Authorized reference method for detection of SARS-CoV-2. Samples were collected at four different geographically distinct sites (one in Urbana, IL; one in Madison, WI; two in Chicago, IL). All nasal swab samples were analyzed in the clinical pathology laboratory at the University of Illinois Chicago Hospital using the Abbott RealTime SARS-CoV-2 assay performed on the Abbott m2000 System with a LoD of 2,700 NDU/mL, and all saliva samples were analyzed using the covidSHIELD assay at the University of Illinois Urbana-Champaign CLIA-registered laboratory. Participants were all over the age of 18 and suspected to have COVID-19 (either with symptoms or with known exposures to someone positive for COVID-19). Of the 120 sample sets that were collected, 31 of the nasal samples were found to be positive and 89 of the nasal samples were found to be negative (Fig. 1e, Extended Data Table 4). Excellent overall concordance (98.3%, 95% CI, 94.1-99.8%), positive percent agreement (96.8%, 95% CI, 83.2-99.9%) and negative percent agreement (98.9%, 95% CI, 93.9-99.9%) were observed for the covidSHIELD assay on the contemporaneously collected saliva samples (for more detailed information on the clinical study, see Methods and Extended Data Tables 5 and 6).

An additional clinical study was performed with 17 individuals confirmed by mid-turbinate nasal swabs analyzed at the Johns Hopkins University School of Medicine to be positive for COVID-19 and to have low viral loads (Ct = 32-42, avg 37). Contemporaneously collected saliva samples were then analyzed using the covidSHIELD assay at the University of Illinois Urbana-Champaign CLIA-registered laboratory. As shown in Fig. 1f, the covidSHIELD assay detected at least two genes for SARS-CoV-2 in 16/17 of these low viral load samples, and one gene for SARS-CoV-2 was detected in the 17^th^ sample. In 15 out of these 17 samples, the average Ct values for the covidSHIELD assay were lower than that of their matched nasal swabs, and the overall average Ct value for the covidSHIELD assay was significantly lower than that of nasal swab-based based test (Avg Ct for covidSHIELD = 31.82, average Ct for nasal awabs = 37.05, p < 0.0004) (For more detailed information on the clinical study, see Methods and Extended Data Table 7).

The U.S. Food and Drug Administration has granted Emergency Use Authorization to the covidSHIELD test: EUA202555 SUMMARY covidSHIELD Assay (https://www.fda.gov/media/146317/download).

## Tell

It was critical to maximize the speed and effectiveness with which the results of the covidSHIELD assay could be communicated, and to couple these results to participation in desired activities to encourage compliance. To achieve these goals, a multidisciplinary team developed a COVID-19 app called Safer Illinois. The app automatically receives the results of the SARS-CoV-2 tests performed at the UIUC CLIA-registered Lab in a manner that is privacy-preserving and HIPAA compliant. The app also displays a cover screen that grants or denies access to campus buildings based on an individual’s most recent covidSHIELD test results (Fig. 1g). A proximity-based and thus privacy-preserving exposure notification feature warns users when they have been in significant contact with someone who has recently tested positive. In order to maintain “Building Access Granted” status, an individual must be up to date on their required testing frequency, not have recently tested positive for COVID-19, and not have a recent exposure notification from the app. The Building Access feature is a key driver of compliance with the testing protocol. The code for Safer Illinois was made publicly available and independently audited by three different independent expert groups. Survey results and online town hall style meetings showed that in general the privacy-preserving features of the app were well-appreciated, but there were still some concerns about privacy and potential for data sharing. Thus, we decided not to mandate the use of Safer Illinois. People could alternatively choose to receive their covidSHIELD test results via a secure email through the campus’ McKinley Health Center, and/or utilize a web-based process to communicate their building entry status.

## Deployment during Fall 2020

After an encouraging pilot study in July 2020, we deployed the complete SHIELD: Target, Test, and Tell platform across our entire community of undergraduate and graduate students, postdoctoral associates, staff and faculty, a population of about 50,000 people, from the time our undergraduates returned on August 15, 2020 until the last scheduled day of the semester on December 23, 2020. Participation in the SHIELD program was required for all students living in Champaign-Urbana, and for all faculty and staff that chose to access campus buildings. We estimate that ∼60% of our students were highly compliant with the required testing throughout the semester, most of the remainder were somewhat compliant, and there was a minority of students who were non-compliant. As summarized in Fig. 2a, during this time we performed >1,000,000 covidSHIELD tests with an average turnaround time of 11.2 hours, and we kept classrooms, research laboratories, and many other University activities open. There were more than 49,000 unique users of Safer Illinois (approximately 94% of the individuals who were eligible to be tested). Over the semester, 94% of the virus test results were transmitted via the app. There were 3.95 million Safer Illinois app sessions (IOS users, 77%; Android users, 23%), more than 920,000 views of the Building Entry Status, 166,000 views of Testing Locations, 26,600 views of Health Guidelines, 25,000 views of the Health Care Team page, and 1,160 digital exposure notifications.

During this time period, our new daily COVID-19 case positivity rates were generally less than 0.5%, we had no evidence of spread within our classrooms or research laboratories, and we also had zero COVID-19-related hospitalizations or deaths among our campus community.

Although in general our positivity rates remained low, there were four periods during the Fall semester during which we observed transient increases in the number of daily cases (Periods I-IV indicated in Fig. 2a). The most notable example occurred at the beginning of our semester (Period I). Based on our modeling we expected that several hundred of our ∼35,000 undergraduate students would be infected with SARS-CoV-2 when they returned to our campus in mid-August 2020 (see Methods). The university required students to test as soon as they arrived on campus, and they were not permitted access to any campus buildings until they received a negative test result. From August 12 – August 23, we conducted a total of 55,034 tests and detected 288 new cases of COVID-19, yielding a new case-positivity rate of 0.52%. During the following week (August 24 – September 21), our covidSHIELD testing revealed a spike in cases (Fig. 2a). Fortunately, because we were testing everyone in our community twice per week, we had an early warning signal and comprehensive dataset that allowed us to respond in a data-driven manner. More than 95% of the new positive cases were in undergraduates, and we identified several clusters in buildings where social activities inconsistent with campus recommendations had been reported. Contact tracing combined with covidSHIELD testing data revealed early signs of potential outbreaks in these buildings.

Guided by these data, we did three things: First, we required that all undergraduate students engage only in essential activities for two weeks. Essential activities were defined as in-person classes, laboratory activities, employment responsibilities, solo outdoor exercise, religious activities and grocery shopping. Second, we modified our testing protocols to prioritize fast and frequent testing of undergraduates, especially those at highest risk of transmitting COVID-19. Undergraduates living in buildings with high numbers of positive cases were required to test three times per week, all other undergraduates continued to be required to test twice per week, and all faculty, staff and graduate students were switched to once per week. Third, we increased the speed with which undergraduates who tested positive were safely isolated using text messaging. While we can not determine the specific impact of these interventions, our observation was that over the course of the next two weeks, our daily case positivity rate dropped from a peak of 2.86% on August 30 to 0.25% on September 12 (Fig. 2a).

For the remainder of the semester, we continued to perform the same approach focused on risk-prioritized fast/frequent testing and rapid isolation. All three of the subsequent increases in cases were similarly followed by rapid declines (Periods II-IV). Period IV is also notable. Classes were pre-scheduled to be moved online after Thanksgiving break, but student surveys revealed that 60% of undergraduates planned to return to Champaign-Urbana after the Thanksgiving break. We expected an increase in cases stemming from travel and holiday gatherings, and we were concerned about social activities during the cold month of December. Thus, all undergraduate students that chose to return to Champaign-Urbana were required to test three times per week. During this time, we also returned faculty, staff and graduate students to a twice-per-week schedule. After a small spike upon the students’ return, our case positivity quickly reduced and generally remained low, reaching 0.25% as we passed the 1,000,000 test mark on December 15. We ended the semester on December 23 with a case positivity rate of 0.16%.

## Analysis of viral load

Because we were testing people in our campus community once, twice, or three times per week, by the time we reached September 2 we had documented negative tests for most of our campus community. This indicated that people testing newly positive were generally still in the early stages of infection. Prior studies suggest that Ct values of greater than 25 (corresponding to lower viral loads) are correlated with reduced ability to recover infectious virus and hence individuals with Ct >25 pose a lower risk for transmission.^10, 27–31^ For the people that first tested positive with the covidSHIELD assay from September 2 – December 23, 73% had recorded Ct values that averaged >25, and the average Ct value was even higher when the most recent negative test occurred within the past few days (Fig. 2b). The covidSHIELD assay was thus likely detecting many of these new positive cases *prior* to infectiousness.

## Head-to-head daily testing with covidSHIELD and an antigen-based lateral flow assay

We also compared the covidSHIELD assay to a lateral flow assay/antigen test for COVID-19. Specifically, a subset of participants in the Division of Intercollegiate Athletics (n=190) were tested every day with both the covidSHIELD saliva RT-qPCR assay and the Quidel Sofia 2 SARS-CoV-2 MT swab Antigen FIA assay from October 1, 2020 to December 23, 2020, for a total of 13,299 comtemporaneous tests. This real world daily testing of asymptomatic people enabled comparison of these two testing methods for their ability to identify infected individuals and remove them from the population in the early stages of infection. As shown in Fig. 2c, this study found six SARS-CoV-2 positive participants during the entire period. All six were identified as positive via the covidSHIELD assay and confirmed to be positive by a subsequent positive covidSHIELD test within the following 4 days. None of these individuals tested positive on the contemporaneously performed lateral flow assays (Fig. 2c). The initial average Ct value for the covidSHIELD test in these 6 individuals was 30.2 +/- 4.73 (Fig. 2d), consistent with the findings in Fig. 2b.

## Analysis of mitigation of SARS-CoV-2 transmission

To probe the extent to which SHIELD reduced transmission on campus, we estimated the 7 day averaged time-dependent reproduction number (R_t_), for the four time periods (**I**-**IV** in Fig. 2a) during which we had observed transient increases in cases of COVID-19 (Fig. 3a-d). The case numbers fell exponentially in time following each of these episodes. The decay rate *g* was measured and used to determine the effective reproduction number R_t_ during these times (top panels).^32^ In addition, R_t_ was alternatively determined using the method of Cori *et al*.^33^ (bottom panels). These estimates are broadly consistent and demonstrate that R_t_ reached as low as 0.5, and was frequently around 0.8-0.9.

We also sought to quantitatively assess the extent to which cases within the campus community influenced or were influenced by each other, and/or by cases in the surrounding community, by examining correlations between sub-populations of the university and surrounding communities. Such an analysis can only reveal the average trends and correlations, and cannot rule out specific instances of transmission that do not conform to the trends. As shown in Fig. 3e, the number of 7-day averaged daily new cases of faculty/staff strongly correlated with that of residents in Champaign County, especially after October 18 (Pearson correlation coefficient 0.86, p-value 7.99×10^-38^, Extended Data Fig. 6a). At the beginning of the semester (around August 31) when there was a short duration spike in daily new cases from undergraduate students, it had little influence on the faculty/staff (Fig. 3f) or on the surrounding Champaign County community (Fig. 3g). Later in the semester (after October 18), as the number of positive cases in Champaign county and faculty/staff increased, the number of cases of undergraduate students followed similar trends (Fig. 3f and 3g; Pearson correlation coefficient for number of cases between undergraduates and faculty/staff = 0.88, p-value 1.33×10^-21^, Extended Data Fig. 6b). All of these data, and additional data from time correlations of residents of Champaign County, undergraduates and faculty/staff (Extended Data Fig. 6) indicate that both faculty/staff and Champaign County cases were essentially uncorrelated with the undergraduate students. Thus the campus population did not drive cases within the surrounding community.

## Other communities with or without SHIELD

To further evaluate the efficacy of the SHIELD program, we compared our results to those observed by other communities operating with or without fast/frequent testing with the covidSHIELD assay.

We first looked at other communities that started using SHIELD sometime in the Winter or Spring of 2020/2021. As shown in Fig. 4a, another university initiated the use of SHIELD at the beginning of the Spring semester in mid-January 2021. The observed initial 7 day average positivity rate for this community was >0.7%. This was rapidly reduced over the period of one month after the introduction of SHIELD, and the positivity rate reached <0.1% by the end of the Spring 2021 semester.

SHIELD testing was similarly introduced in the middle of the Spring semester at a pair of high schools that required masking and social distancing for the entire semester (Fig. 4b). The observed 7 day average positivity rate was initially found to be ∼0.5%, and after the introduction of SHIELD, this was quickly reduced to and maintained for the rest of the semester at </= 0.1%. This provided an opportunity to observe the positivity rate before and after the impact of SHIELD. Similar results were observed at a federal courthouse and a large corporate campus, and again masking and social distancing were in place before and after the introduction of SHIELD (Fig. 4c and d). Our saliva-processing protocol now forms the basis for testing the populations of >75 colleges and universities (for a partial list see Extended Data Fig. 7), and it is also being used to provide frequent testing for multiple K-12 schools, companies, and municipalities in the United States and abroad.

Finally, we compared predicted and observed COVID-19 mortality among counties with Big 10 Universities (n=13) from July 6 to December 23, 2020. This allowed us to compare the outcomes for UIUC’s surrounding Champaign County to those of the other BIG 10 Universities, all of which had similar requirements for masking and social distancing, but none of which had the SHIELD program. To account for the potential effects of socio-economic disparities and the demographic makeup of a given county, the overall mean COVID-19 mortality rate in each county was race- and age-adjusted and assessed as a function of social vulnerability using linear regression (Fig. 4e). The social vulnerability index is used as a proxy to determine a community’s ability to prevent human suffering and financial hardships in the event of a disaster.^36^ We also controlled our analysis by state, given the widely varying policy factors that influence mortality rates (e.g. mask mandates, business closings, etc.). Using a best-fit model for all Big 10 University counties, Champaign was the only county to observe statistically significantly lower COVID-19 mortality than predicted, reducing mortality by 14.6% (95% CI 9.89, 19.25; Fig. 3e). This analysis provides strong evidence that the SHIELD Target, Test, and Tell program uniquely resulted in a protective effect for the communities in Champaign County.

There are three important contributions of this work, the first of which is the development and deployment of the multimodal “SHIELD: Target, Test, and Tell” program; a comprehensive effort designed to mitigate the spread of COVID-19 at a large public university, prevented community transmission, and allowed for the continuation of in-person classes amidst the pandemic. The second contribution is the description of the novel, scalable, saliva-based RT-qPCR assay for SARS-CoV-2 that bypasses RNA extraction. The third contribution is the evidence that fast, frequent testing can mitigate the transmission of the SARS-CoV-2 virus after spikes in infections occur.

Combined with masking and social distancing, fast/frequent testing helped mitigate the spread of COVID-19 in the Fall 2020 semester for a large and diverse public University community. Even though our classes, laboratories and local businesses stayed open, we found no evidence of transmission from students to faculty or staff, no evidence of transmission from the university community to the surrounding community, and no one in our campus community became seriously ill or died as a result of COVID-19 during this timeframe. The covidSHIELD saliva test is highly scalable and easy to implement, allowing fast, frequent and accurate testing in a large community. Our data suggests that every other day testing is able to identify individuals at earlier stages of infections as indicated by lower average viral loads, and daily testing is even better. While daily testing may not be practical on scale, such high frequency testing may be appropriate for certain at-risk groups, including those in congregate living and high contact athletics. The covidSHIELD assay and lessons learned from our experience in the Fall of 2020 helped other communities achieve similar outcomes in the Winter of 2020/Spring of 2021. Our results stand to better enable communities to open more safely as the world bridges to widespread vaccination against COVID-19. They also provide a playbook for addressing the next pandemic.

## Methods

### Agent Based Modeling

We simulate the spread of the epidemic using a full-scale stochastic agent-based model (ABM), which tracks the movement and disease stage of a large number of individuals as they attend both academic functions (like classes, libraries, and study groups) and social events (including eating at restaurants, congregating at bars, and attending parties). The core of the model comprises a set of 46,850 individuals who each follow independent schedules, which specify how the agents should move between different physical locations or *zones.* When an agent enters a zone, a random position in the zone is selected and the agent is assumed to stay at this position until he/she leaves the zone.

The model base is augmented by an infection model, which defines how agents become infected and infect other agents. Supplemental modules for testing, contact tracing, and quarantine/isolation can be interfaced with the core model to compare the effectiveness of various mitigation strategies.

#### A. Infection model description

Motivated by the airborne transmission dynamics of COVID-19, we have adopted the concept of “infection quantum”, which is defined as the dose of airborne droplet nuclei required to cause infection in 63% of susceptible persons.^37^ We introduce a hybrid transmission model which decomposes the infectious droplets into two parts: (1) large droplets with sizes larger than 10 microns which stay within a circle of radius 2-meter from an infected agent and only infect its neighbors within the circle; and (2) small airborne droplet nuclei with dimensions less than 10 microns which spread homogeneously over the zone. Accumulating more quanta corresponds to increasing the probability of being infected.

When an agent leaves a zone, they are infected with a probability that depends on the total number of quanta they accumulated in that zone *N = ∫ndt*. Specifically, the infection probability is given by:

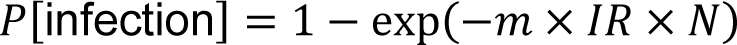

where *m* is a modifier that accounts for modulation of inhaled quanta due to factors such as mask wearing, and *IR* is the inhalation rate of the susceptible individual.^37^

The quanta emission rate from an infectious agent depends on their activities in different zones. For example, since students mostly stay silent during classes, the emission rate is estimated to be 4 quanta per hour. As a comparison, since agents often speak loudly in bars/parties, the quanta emission rate is assumed to be 150 quanta per hour. The emission rates are derived from Linsey *et al.*^38^. In addition, the quanta emission rate is proportional to the infectiousness of the infected agent which vary with the in-host viral dynamics. The infectiousness starts to be non-zero after 2 days from the time exposure. This is mean latent period estimated for COVID-19. The infectiousness profile utilized in the model is obtained from a within-host model for COVID-19 that was calibrated by measurements of viral load dynamics.^13^

The droplets exhaled by infected agents containing infectious viral particles are coarsely divided into two components based on the size of droplets: small airborne droplets (≤ 10μ*m* in diameter) and large droplets (> 10μ*m* in diameter). The small airborne droplets can suspend in the air for long periods of time due to the limited influence of gravity.^39^As a result, airborne droplets are heavily influenced by the ventilation rate of the zone and a high ventilation rate can effectively reduce the transmission risk as will be discussed shortly. Since the small droplets spread over the entire zone (depends on the volume of the zone), viruses within exhaled infected agents can infect susceptible agents far away from infectors, and thus lead to long-range infections. The large droplets fall quickly to the ground due to gravity and thus spread only over a limited distance away from the infectious agent (typically less than 2 meters), making only short-range infections possible. The ratio of infectious viral particles between large droplets and airborne droplets *f* is adjusted to reproduce an observed initial doubling time of unmitigated epidemic equal to 2.5 days. This doubling time is motivated by observations of growth rate in COVID-19 cases in Chicago during February/early March 2020. The value of *f* used in this study is equal to 3.

Viral quanta cumulated from either large or small droplets are denoted as *n*_short-range_(*t*) and *n*_long-range_(*t*) respectively. Small droplets spread over a longer distance and are eventually well-mixed over the zone modulated by the ventilation:

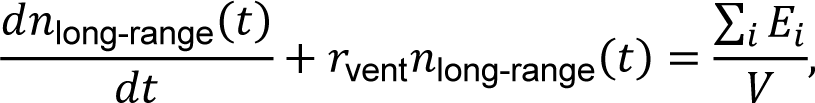

where *r*_vent_ is the ventilation rate, *i* loops over all infected agents, and *V* is the volume of the zone.

Large droplets follow equations for short-range transmission dynamics modulated by both ventilation and gravity:

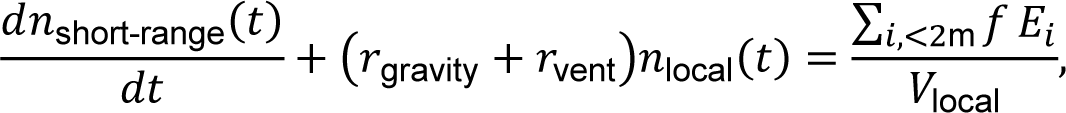

where *r*_gravity_ is the inverse of mean duration that large droplets stay in the air, *i* loops over all infected agents within 2-meter range of the susceptible, and *V*_short-range_ is the volume of a cylinder with radius 2 meters and height 2 meters. To simplify the computation, *n*_short-range_(*t*) is assumed to be in a steady state and thus 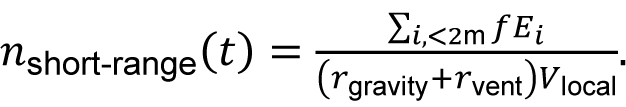. The quanta emission rates for different agents in various zones are described in Extended Data Table 8.

For each susceptible agent, the probability of being infected for each zone is independent. The infection probability for one susceptible in a zone depends on the accumulated quanta *N* and *N* is a time-integral of 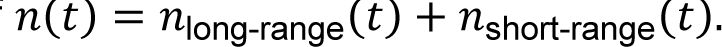.

The disease transmission dynamics in homes/dorm environments is hard to be modeled for two reasons: (1) the unknown co-habitation information of agents and (2) too many independent houses/homes/dorm rooms to be tracked. This causes computational complexity. To consider the disease transmission due to cohabitation at homes/dorms, all agents in the simulation are grouped into pairs and they are roommates to each other. The disease transmission can happen between two roommates and the probability is computed from the quanta emission rate for breathing, the volume in typical dormitories, and the ventilation rate in dorms. For each night, an infection event between one susceptible roommate and the other infected roommate is probabilistic.

#### B. Schedules

Agents’ schedules are generated before running the simulation and are motivated by real information about both undergraduate and graduate students’ class schedules of the semester Fall 2019 as provided by the University of Illinois at Urbana-Champaign. A schedule comprises a list of zones and times that define where an agent should be as a function of time. To adapt for the agent-based model, the duration of classes is assumed to have a granularity of 30 minutes. Zones are divided into groups according to their function, and may be associated with academic courses, libraries or study groups, cafes, restaurants, or bars/parties. Each zone has a defined, static size and is associated with certain quanta emission rates (see Extended Data Table 8), e.g., the emission of quanta is higher in bars than classrooms since one is more likely to be talking in a bar. Besides the student population, there are also non-student agents (1 professor for every class and 5 workers in every bar, restaurant, cafe, or library). These other agents are assumed to stay at home for most of time except going to their working zones. There are assumed to be 15,000 students living in 300 dormitories, while the remaining students are divided into the other home zones. For simplicity, 300 dormitories are assumed to be the same size, with 50 students living in each one. The total number of non-student agents is 2446.

The disease transmission at home/dorm is assumed to be stochastic and it occurs between randomly paired roommates with a constant probability that is inferred from extended exposure overnight constrained by the typical volume and the ventilation rate of dormitories.

Although our assignment of courses is based on a real schedule, we can only synthesize the agents’ out-of-school schedules. We use the following model to fill in the blanks:

- Agents are assumed to be home/dorm between 21h00 and 8h00.
- Agents do not skip classes.
- Agents eat at restaurants twice a day for lunch and dinner between (10h-14h) and (17h and 21h) respectively. An agent will spend a random amount of time at the restaurant or dining home:

- Lunch: uniform random between 0.5h to 1h.
- Dinner: uniform random between 0.5h to 2h.
- Agents who stay at home/dorm, in the library, at a cafe, or go to bars tend to stay a long time and the assumed time interval spent is given in Extended Data Table 9.
- 7000 students would go to bars after 6p.m., and agents who go to bars/parties only do so on the Thursdays and weekends. (Agents may stay overnight in bars).
- On a daily basis:

- 2500 agents would go to cafes if they are free at the time.
- 11250 students would go to libraries or congregate in study groups if they are free at the time.
- 5 out of 50 students living in each dormitory would go to a dorm party each night for 2 hours.
- Agents would stay at home/dorm for a while if they have nothing else to do at the time.

To simulate the real-world campus, the schedule also added 2 groups of specialized agents: professors/staff and workers. Each course section is assigned with a course staff who would teach that section and stay at that class zone during class time. Each non-class zone (except home/dorm zone and lab zone) would have 5 workers who would stay at their working zone from 8 a.m. to 9 p.m. Workers will go for lunches and dinners as everyone else and this provides an opportunity for mixing between students and non-student agents.

In addition to the above specified in-semester schedules of all agents, we have also incorporated a pre-semester schedule that reflect student activities during the move-in week before the start of the semester. This is crucial because some agents may come to campus already infected and hence they will be detected by testing during initial screening and may also be capable of infecting other susceptible individuals while the mitigation strategies (such as mask-wearing and hybrid classes) are not fully effective yet. We have summarized our calculation for entry screening in an earlier report.^40^ Given the prevalence of COVID-19 in Illinois in August 2020, we estimated that approximately 300 infected agents will be detected in entry screening.^40^ In the 3-day pre-semester period, agents spend more time in restaurants, cafes, and bars. All agents are required to receive a universal testing screening in the next 2 days upon campus arrival.

#### C. Quarantine and Isolation

Agents who are test positive will have their regular schedules modified through mandatory isolation and thus will be disconnected from the general population. Potentially exposed individuals are also identified through contact tracing and may be disconnected from the general population through mandatory quarantine. Thus, in our model, individuals may be disconnected from the population due to two reasons: (1) they have been confirmed as infected by testing. We call them: *isolated*. or (2) they have not been confirmed yet but they might have been exposed to an infectious individual and are identified through contact tracing. We call them *quarantined*.

In the real world, quarantine can happen for a variety of reasons including either self-quarantine (e.g., following self-identification of symptoms) or as the result of manual contact tracing or exposure notification (where someone is directed to isolate because they have recently been in contact with an infected individual). In our model, to consider the worst-case scenario, we ignore the case of self-quarantine and attribute the quarantine to the manual contact tracing or exposure notification following an index case (i.e. a positive case). Individuals are quarantined following the manual contact tracing procedure from CDC and exposure notification procedure specified in the Safer Illinois exposure notification app utilized in UIUC. Individuals who are identified as close contacts of an index by manual contact tracers are quarantined for 14 days. If they show any symptoms during the 14-day quarantine period and are tested positive, they will be switched to an isolation protocol. Besides, individuals may be notified by the exposure notification app if their risk scores are larger than a certain threshold. Those individuals notified by the app will be quarantined for 5 days before receiving a test.

#### D. Testing

The testing module enables agents to be checked for the disease and be isolated from the general population if the testing procedure identifies them as having been infected. Our implementation of testing makes several assumptions:

`o` Testing is only available for part of the day (between 8 am and 6 pm).

`o` An infected agent may be incorrectly tested negative. For this modeling exercise, the false negative rate was assumed to be 11.1% based on initial studies.^23^ The false negative rate is likely much lower: In the clinical study shown in Fig. 1d, negative percent agreement for covidSHIELD assay with nasal swab-based assay was 98.9% (95% CI, 93.9-99.9%).

`o` The false positive rate of the test was assumed to be zero.

`o` The delay between having been tested and receiving the results of the test is 5 hours. The testing results of all collected testing samples will be notified as soon as possible without the working hour limit.

#### E. Contact tracing and exposure notification

We implement modules for both manual contact tracing and automatic app-based exposure notification. The manual contact tracing is performed by manual contact tracers. Once an index case is identified by the test, the manual contact tracing starts to find the index’s close contacts (i.e., contacts whose distance to the index is less than 2 meters) staying more than 15 minutes with the index case. Given the fact that such a manual contact tracing requires a large force of manual contact tracers and not every close contact cannot be accurately identified, here we assume that only 50% of close contacts may be successfully traced. Close contacts in classes are hard to be identified due to random mixing that may happen between students. As a result, the whole class will be moved online and all agents in this class are quarantined once an infected case is diagnosed in this class.

The exposure notification (EN) relies on the Privacy-Preserving Contact Tracing Project developed by Apple and Google. It is a decentralized protocol that combines the Bluetooth technology with the privacy-preserving cryptography. For example, if two agents that have installed the exposure notification app such as Safer Illinois developed locally in UIUC, two phones with Bluetooth opened constantly send encrypted keys to nearby phones having the same app via Bluetooth. If the distance within two phones is less than 2 meters, encrypted keys of both phones are exchanged and saved locally on the phones. All saved keys older than 14 days will be deleted. When a user who is tested positive sends the positive test result to the central server, the encrypted key of this confirmed case will be uploaded to the central server. The encrypted keys of confirmed cases will be automatically downloaded via the app by all users and they are compared to the saved keys locally. Whenever there is a match between the encrypted keys of confirmed cases and locally saved keys, the risk score of the user will be updated. If the total risk score of the user in the past 14 days is larger than a threshold, the user will be notified by the exposure notification app.

We enhance the exposure notification through a risk-weighted approach. The probability of being infected depends on three factors: infectiousness of the index, contact duration, and the zone risk. Thus, the risk-weighted protocol is developed here to reflect these factors in the risk score. The risk score is defined as contact duration (in hours) with all index cases weighted by the infectiousness of all index cases. The infectiousness is scaled to make the peak infectiousness as 1. Given the probability of being infected in zones such as bars is much higher than the infection probabilities in all other zones, the risk score in such risky zones is increased to reflect the true process of disease transmission. To achieve this, all agents’ activities during weekends’ and Thursdays’ nights will be treated as in bars and their risk scores are multiplied by 10. The notification happens when the total risk score passes 2.

#### F. Additional mitigation strategies

In addition to the testing, contact tracing, and exposure notification, there are several extra mitigation strategies effective for reducing the infection:

`o` Hybrid classes: a fraction of classes (especially large classes) is moved online to reduce the contact hours between students. In the model, all classes with the size over 50 are moved online and all students are assumed to stay at home/dorm when they are taking the online class.

`o` Mask-wearing: masks can reduce both the emission rate and the inhaled rate of viral quanta. In our model, the overall reduction coefficient for the mask-wearing is assumed to be 50% when both the infector and infectee wear masks. The mask efficiency in reducing emission is assumed to be 30%. The mask efficiency in reducing inhalation is assumed to be 30%. These numbers are conservatively assumed consistent with a single layer cloth mask. Thus, if either the infector or the susceptible individuals does not wear mask, the mask reduction in transmission is only 30%. If both agents wear masks, the effect of masking is quadratic and the transmission is reduced by (1-0.3)^2^ = .49, or approximately ∼ 50%

### Evaluating the multiplier, M, for testing frequency

We adopted the following infectiousness profile, after Goyal et al. 2020^13^, which includes 2-day latency period of [0.0, 0.0, 0.148, 1.0, 0.823, 0.426, 0.202, 0.078, 0.042, 0.057, 0.009, 0.0, 0.0, 0.0, 0.0, 0.0, 0.0]. The sum of this infectiousness list is therefore 2.785. The effect of testing frequency on reducing the transmission chain may be estimated as follows:

A. Testing once a week: With one test a week, agents may test positive and get isolated on days 3, 4, 5, 6, 7, 8 or 9 after exposure with a probability = 1/7. Therefore, the expected sum of infectiousness profile modulated by 1 test a week is (1/7) * 0.148 + (1/7) * (0.148 + 1.0) + (1/7) * (0.148+1.0+0.823) + (1/7) * (0.148+1.0+0.823+0.426) + (1/7) * (0.148+1.0+0.823+0.426+0.202) + (1/7) * (0.148+1.0+0.823+0.426+0.202+0.078) + (1/7) * (0.148+1.0+0.823+0.426+0.202+0.078+0.042) = 1.951. This generates a **R0 multiplier = 1.951 / 2.785 = 70.5%**.
B. Testing two times a week: With two tests a week, agents may test positive and get isolated on days 3, 4, or 5 after exposure with a probability = 2/7, while agents may test positive and get isolated 5.5 days after exposure with a probability = 1/7. Therefore, the expected sum of infectiousness profile modulated by 2 tests a week is (2/7) * 0.148+ (2/7) * (0.148 + 1.0) + (2/7) * (0.148+1.0+0.823) + (1/7) * (0.148+1.0+0.823+0.426/2) = 1.245, generating a **R0 multiplier = 1.245 / 2.785 = 44.7%**

### Computation of R_0_ in the hybrid transmission model

In the hybrid transmission model, the infection occurs through the accumulation of infectious quanta emitted from all infectious agents present in the zone. In other words, the concept of transmission pair is not clear and instead, each infectious agent has a fractional contribution to an infection event. As a result, here we attempt to estimate R_0_ as a sum of fractional contributions. First, for each infection event that occurred in the simulation, we recorded the infection zone and time for this individual and obtained all infectious agents within the zone at that time according to the schedule and excluded all isolated agents. Second, for an infection event with n infectors, we assigned a fractional R_0_ contribution 1/n for each infector. Finally, during the early period of the epidemic, the fractional R_0_ for all infection events are assigned to all infectious agents in presence, and R_0_ for each infected agent is a sum of all fractional R_0_.

### Acquisition and processing of clinical samples

All clinical samples from study participants were collected in accordance with Western IRB-approved protocol number 20203538. Participants were recruited from populations seeking SARS-CoV-2 tests and were included if they 1) reported symptoms consistent with COVID-19 or suspected exposure to an infected individual, 2) had never tested positive for SARS-CoV-2, 3) were at least 18 years of age, and 4) spoke English. All participants provided informed consent at the time of recruitment. Participants provided a 2 mL saliva sample, and a health professional collected a nasopharyngeal swab following standard procedures. The saliva sample was transported to the Veterinary Diagnostic Laboratory within 24 hours for analysis following the outlined procedure. The nasopharyngeal swab was inserted into viral transport media and stored at −80°C until analysis with an FDA-approved comparator at an independent diagnostic laboratory.

Extended Data Table 4 summarizes the method comparison study completed to support the correlation between saliva samples processed with covidSHIELD and nasal samples processed with Abbott RealTime SARS-CoV-2 assay performed on the Abbott m2000 System. Extended Data Table 5 outlines the details to capture on case report form. Extended Data Table 6 summarizes the 26 clinical samples were split into two aliquots upon receipt, one set was processed using our covidSHIELD assay and the other set was subjected to RNA extraction using MagMax Viral/Pathogen II (MVP II) Nucleic Acid Isolation Kit (ThermoFisher). 5uL of either processed saliva (in 1:1 2x TBE/Tween-20 buffer) or purified RNA (in water) were subsequently used as templates for RT-qPCR. Extended Data Table 7 summarizes the comparative study of mid-turbinate (MT) swab and saliva from 17 individuals identified with low viral load based on MT swab analyzed using Abbott Alinity RT-PCR at the Johns Hopkins University School of Medicine. Contemporaneously collected saliva samples from the same individuals were analyzed using the covidSHIELD assay at the University of Illinois Urbana-Champaign CLIA-registered laboratory.

### SARS-CoV-2 inactivated virus

In most experiments, fresh pooled saliva were spiked with gamma-irradiated (BEI cat# NR-52287, Lot no. 70033322) SARS-CoV-2 virions. SARS-Related Coronavirus 2, Isolate USA-WA1/2020, Gamma-irradiated, NR-52287 was deposited by the Centers for Disease Control and Prevention and obtained through BEI Resources, NIAID, NIH. The reported genome copy number pre-inactivation for γ-irradiated SARS-CoV-2 is 1.7×10^9^ genome equivalents/mL for the specified lot number. All virus stocks were aliquoted in small volumes and stored at −80°C. Stocks were serially diluted to the correct concentration in RNase-free water on the day of experimentation.

### Collection and processing of fresh saliva from healthy donors for Limit of Detection (LoD) assay

Fresh saliva was collected from healthy individuals in 50 mL conical tubes (BD Falcon) in accordance with University of Illinois at Urbana-Champaign IBC-approved protocol numbers 4604 and 4589. Known amounts of the SARS-CoV-2 inactivated virus (BEI) were spiked into saliva samples. Samples were processed according to the covidSHIELD instructions for use (https://www.fda.gov/media/146317/download). Briefly, samples were incubated in a hot water bath at 95°C for 30 min. After cooling the sample on ice, 100 uL saliva was transferred to 96-deep-well plates pre-loaded with 100 uL of 2x TBE + 1% Tween-20 buffer at 1:1 dilution ratio. 5uL of this sample preparation was used as template for RT-qPCR reactions.

We performed a multiplex RT-qPCR assay using the TaqPath RT-PCR COVID-19 kit (Thermo Fisher CN A47814) together with the TaqPath 1-step master mix – No ROX (Thermo Fisher CN A28523). All RT-qPCR reactions, comprised of 5uL template + 5uL of reaction mix (2.5uL TaqPath 1-step master mix, 0.5uL TaqPath primer/probe mix, 1.0uL MS2, and 1.0 rnase-free water), were performed in 384-well reaction plates in a QuantStudio 7 system (Applied Biosciences). The RT-qPCR was run using the standard mode, consisting of a hold stage at 25°C for 2 min, 53°C for 10 min, and 95°C for 2 min, followed by 40 cycles of a PCR stage at 95°C for 3 sec then 60°C for 30 sec; with a 1.6°C/sec ramp up and ramp down rate. The limit of detection (LoD) of the assay was performed by serial dilution of γ-irradiated SARS-CoV-2 (0-5.0×10^5^ viral copies/mL) used to spike pooled fresh saliva samples. LoD experiments were repeatedly performed at least five times in different machines.

In some experiments, the CDC-approved assay was used to validate our data using the TaqPath 1-step mix (Thermo Fisher CN A15300). Primers and probes targeting the N1, N2, and RP genes were purchased from Integrated DNA Technologies as listed: nCOV_N1 Forward Primer Aliquot (CN 10006830), nCOV_N1 Reverse Primer Aliquot (CN 10006831), nCOV_N1 Probe Aliquot (CN 10006832), nCOV_N2 Forward Primer Aliquot (CN 10006833), nCOV_N2 Reverse Primer Aliquot (CN 10006834), nCOV_N2 Probe Aliquot (CN 10006835), RNase P Forward Primer Aliquot (CN 10006836), RNase P Reverse Primer Aliquot (CN 10006837), RNase P Probe Aliquot (CN 10006838). The 2019-nCoV_N_Positive Control (IDT CN 10006625) was used as positive control at 50 copies/μL dilution. LoD experiments using CDC primers were performed at least three times.

### RT-qPCR Data analysis

Following completion of RT-qPCR, data were processed using QuantStudio Design and Analysis Software (version 2.4.3) with a threshold setting of 10,000 and a baseline setting of 5. Cycle threshold (Ct) cut-off was set at 39. Ct values were plotted as single replicate values on a scatter plot, using GraphPad Prism 8 (version 8.4.2).

### SHIELD deployment in other communities

Saliva samples were collected from a large public university, a community college, a pair of high schools, a federal courthouse, and a large private corporate campus. Samples were processed and analyzed using the covidSHIELD assay (FDA EUA 202555). The test results were evaluated according to the interpretation tables in the EUA Summary (https://www.fda.gov/media/146317/download). The weekly averaged positivity rates are plotted againts the period the samples were processed.

### Development of Safer Illinois app

Prior to the COVID-19 pandemic, a large collaborative effort led by the Safe, Healthy Community Initiative on campus had been developing an open source software platform called Rokwire.^41^ Rokwire is designed to make it easy for individuals and organizations to build apps for mobile devices that support smarter, healthier communities. We had been using the University of Illinois at Urbana-Champaign campus as a test bed for the development of Rokwire — the platform — and the first app built upon it — the Illinois app. Safer Illinois was built on the Rokwire platform and the source code was made open source August 14, 2020.^42^

As part of the process of designing Safer Illinois, we met with and sought feedback from a diverse set of stakeholders: faculty and students, mental health advocates, leadership in Student Affairs, the Faculty Senate, the Graduate Employees Organization, and a range of individuals with expertise in digital privacy. Collectively, these groups expressed a variety of concerns related to privacy and data security.

We took multiple actions to address the concerns expressed. We built privacy into Safer Illinois from the ground up. We made modifications to a beta version of the app to minimize the data we collected and stored so that collected only the data necessary to allow the app to function. We designed the app to store data related to Exposure Notification for the shortest possible period and then delete it. We ensured that users could delete their data at any time from both the app and servers. We made our privacy notice novice friendly, so that all consent language allowed users to understand up front exactly what data we collect, what we do with that data, how long we keep it, and how users can manage their data.^43^

## Code Availability

Source code repository of “Safer Illinois” App - the official COVID-19 app of the University of Illinois: https://github.com/rokwire/safer-illinois-app

## Data Availability

Aggregate case and testing data are publicly available at https://go.illinois.edu/COVIDTestingData All other data may be requested through the COVID Research Oversight Committee at https://forms.illinois.edu/sec/1409755003 Link to the code for analysis of county-level mortality in the BigTen: https://github.com/juel15401/Big10UniversityCounties_COVID.git Code Availability
Source code repository of "Safer Illinois" App - the official COVID-19 app of the University of Illinois: https://github.com/rokwire/safer-illinois-app

## Acknowledgements

We gratefully acknowledge University of Illinois at Urbana-Champaign for funding. UIUC has also received support from CIMIT through the POCTRN program. CIMIT and POCTRN are supported by the RADx Tech program and have been funded in part with federal funds from the National Institute of Biomedical Imaging and Bioengineering, National Institutes of Health, Department of Health and Human Services, under Grant No. U54EB015408. We dedicate this paper to our colleague and co-author Joseph Gregory Gulick who passed away in February 2021. He made critical contributions to the success of the SHIELD program. We would also like to acknowledge Michael Vincent and Diana Yates (Public Affairs, College of Media, University of Illinois at Urbana-Champaign) for editorial support. This work was supported by the University of Illinois System Office, the Office of the Vice-Chancellor for Research and Innovation, the Grainger College of Engineering, and the Department of Physics at the University of Illinois at Urbana-Champaign. Z.J.W. is supported in part by the United States Department of Energy Computational Science Graduate Fellowship, provided under Grant No. DE-FG02-97ER25308. A.E. acknowledges partial support by NSF CAREER Award No. 1753249. This work made use of the Illinois Campus Cluster, a computing resource that is operated by the Illinois Campus Cluster Program (ICCP) in conjunction with the National Center for Supercomputing Applications (NCSA) and which is supported by funds from the University of Illinois at Urbana-Champaign. This research was partially done at, and used resources of the Center for Functional Nanomaterials, which is a U.S. DOE Office of Science Facility, at Brookhaven National Laboratory under Contract No.∼DE-SC0012704. This study was also partially funded by the National Institute for Biomedical Imaging and Bioengineering grant number U54EB027690, part of the National Institutes of Health.

## Author Contributions

M.D.B., P.J.H., T.M.F., R.L.S., N.D.G., W.C.S., J.T.W., S.A.M., A.C.C., R.J.J., and T.L.K. conceived, designed, and guided the research, M.D.B., P.J.H., T.M.F., R.L.S., N.D.G., W.C.S., and D.R.E.R. wrote the paper, M.D.B., P.J.H., T.M.F., N.D.G.,R.L.S., W.C.S., J.T.W., C.B.B., A.V, S.A.M., A.C.C., R.J.J., T.L.K., K.C.W, K.W, R.S.W., E.V., S.V., E.A.T., M.A.T., R.T. M.S., C.S., T.H.S., C.S., J.M.S., A.S., M.E.S., L.N.R., E.R., M.L.R., J.R., J.Q., M.C.P., C.P., J.M.P., J.P., A.P., N.A.P., M.N., A.D.M., K.M., M.M., H.H.M., J.M., A.M., K.G.M., R.M., J.M., Y.C.M., R.L.L., L.M.K., T.K., A.K., P.K., I.J., N.I., A.A.H., A.H., R.M.H., A.G., B.G., D.G., S.A.G., N.G., N.G., B.W.F., D.F., J.F., T.E., A.E., J.E., M.C., M.C., A.C., H.C., J.B., J.B., I.B., J.D.B., K.M.B., M.B., R.L.B., M.J.L., J.E.N., N.J.C., D.L.Y., M.H.L., G.S., J.C.C., M.G., J.S.M., R.B., J.A.V., J.L., W.E.W., D.B.Y., D.S.S., S.P.B., L.M., P.K.A., E.V., M.J., J.L.S., A.C.V., C.H., R.N.K., J.A.P., C.S., J.B., D.M.K., N.W., G.D., C.B-P., B.R.B., L.W-B., M.DL., K.W., K.D., M.R.F.J., J.D.S., C.B., E.S., R.C.P.J., P.M.H., M.P.S., T.J.N., J.G., A.M., I.J.G., S.P.S. J.G.G., N.P.V., J.M.P., S.J.P., Z.L., H.Z., A.V.T., Z.J.W., A.E., S.M., J.U., G.N.W., T.W., R.L.F., L.W., K.J.G., F.G.A., R.L.H., and D.R.E.R. performed the research.

## Competing Interest Declaration

### Financial competing interests

University of Illinois at Urbana-Champaign has filed a patent application related to the saliva-based assay described herein (U.S. Patent Application No.: 63/040,612), and D.R.E.R., R.L.H., F.G.A., K.J.G., L.W., C.R.B., M.D.B., T.M.F., and P.J.H are co-inventors. S.J.P., J.M.P., W.C.S., R.J.J., and A.C.C. are associated with Rokwire, which created the Safer Illinois App. G.D. and N.W. are associated with TekMill which created some of the automation used in the VDL COVID-19 Testing Lab at UIUC.

### Non-financial competing interests

P.J.H., A.C.C., S.A.M. and J.T.W. are Members of the Board of Managers for the University Related Organization SHIELD T3, LLC, which has obtained a non-exclusive license to this technology.

## Extended Data Figure Legends

**Extended Data Figure 1.**
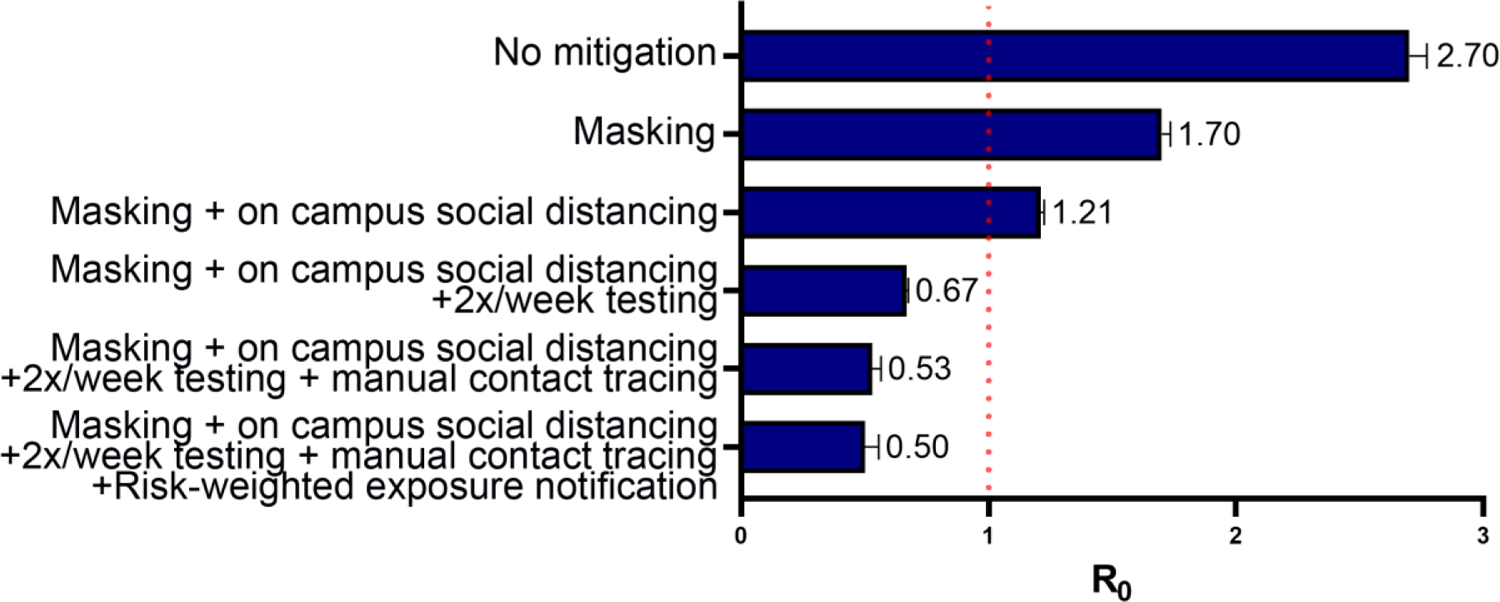
The effect of different mitigation interventions on the basic reproduction number R_0_ as computed in our agent-based model assuming that 60% agents are compliant with testing, isolation and quarantine. If R_0_ is greater than one (orange dashed line at R_0_=1), the epidemic grows exponentially.

**Extended Data Figure 2.**
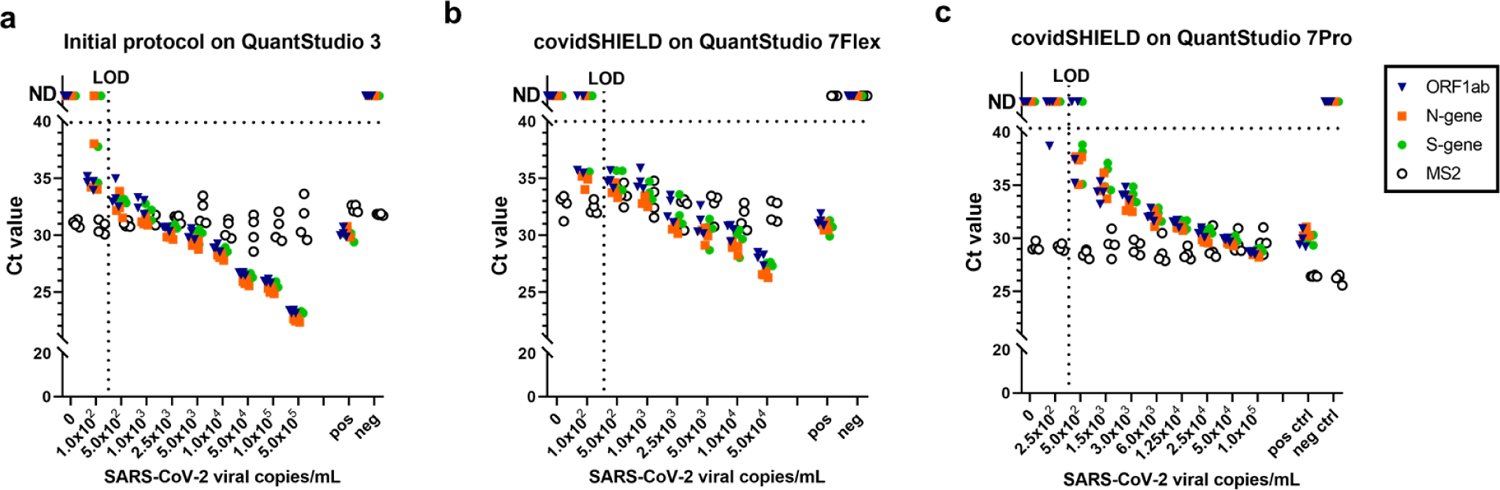
Limit of Detection (LoD) for assessment of SARS-CoV-2 nucleic acid from saliva, comparing the initial protocol performed on QuantStudio 3 **(a)** to the covidSHIELD protocol performed either on QuantStudio 7Flex **(b)** or QuantStudio 7Pro **(c)**. Saliva in 50mL conical tubes was spiked with the indicated amount of gamma-irradiated SARS-CoV-2 prior to heating at 95°C for 30 minutes. Samples were processed using the covidSHIELD assay. PCR plates were run on 3 different QuantStudio models together with a positive control (pos; SARS-CoV-2 positive control, 5.0×10^3^ copies/mL) and a negative control (neg; water). Data in quadruplicates were analyzed for SARS-CoV-2 ORF1ab (blue triangle), N-gene (orange square), and S-gene (green circle), and MS2 (open circle). MS2 bacteriophage was added to the PCR reaction mix as internal control. Undetermined Ct values are plotted as ND. The LoD was set at the lowest concentration at which 2 out of 3 viral target genes were detected. LoD experiments were performed at least five times in different machines.

**Extended Data Figure 3.**
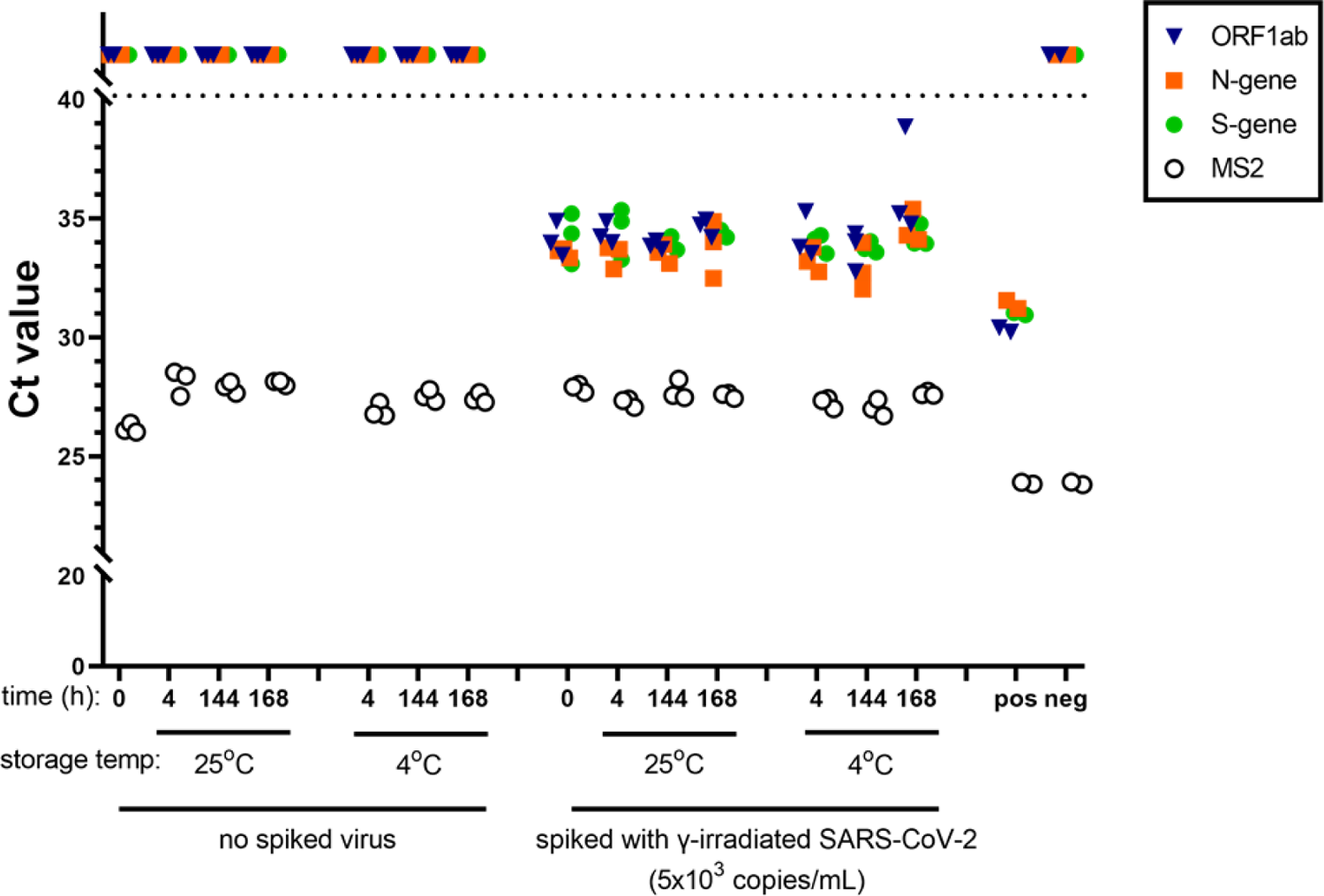
Saliva samples could be stored at room temperature for at least 7 days prior to heating and analysis without loss of sensitivity. Saliva from a SARS-CoV-2 negative subject was collected in 50 mL conical tubes. Sample was divided into sets of aliquots (one set for the negative samples and one for the positive sample). The positive samples were created by spiking the saliva with γ-irradiated SARS-CoV-2 at 5.0×10^3^ viral copies/mL. Samples were further split into smaller groups for storage at either room temperature (25°C) or at 4°C at different time points. Following the incubation period, all samples were processed using the covidSHIELD assay, and together with a positive control (pos; SARS-CoV-2 positive control, 5.0×10^3^ copies/mL) and a negative control (neg; water), were directly analyzed by RT-qPCR in triplicates for SARS-CoV-2 ORF1ab (blue triangle), N-gene (orange square), and S-gene (green circle), and MS2 (open circle). MS2 bacteriophage was added to the PCR reaction mix as internal control. Undetermined Ct values are plotted at 0. Saliva stability experiment prior to heat inactivation was repeated twice.

**Extended Data Figure 4.**
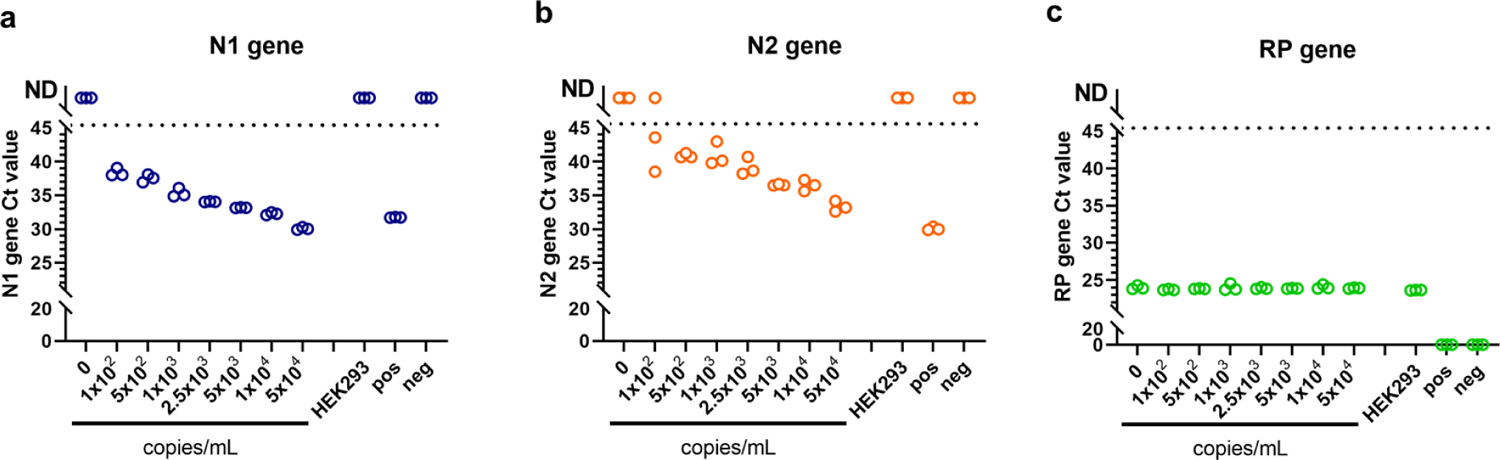
LOD of direct saliva-to-RT-qPCR SARS-CoV-2 nucleic acid detection using CDC-approved primers and probes. γ-irradiated SARS-CoV-2 was spiked into fresh human saliva (SARS-CoV-2 negative) in 1X Tris-Borate-EDTA buffer (TBE) at 1.0×10^2^, 5.0×10^2^, 1.0×10^3^, 2.5×10^3^, 5.0×10^3^, 1.0×10^4^, and 5.0×10^4^ viral copies/mL. Samples were incubated at 95°C for 30 min. Virus-spiked saliva samples, a positive control (pos; SARS-CoV-2 positive control, 5.0×10^3^ copies/mL) and a negative control (neg; water) were directly analyzed by RT-qPCR, in triplicate, for SARS-CoV-2 N1 gene (**a**) and N2 gene (**b**), and the human RP gene (**c**). Undetermined Ct values are plotted at 0. LoD experiments using CDC primers were performed at least three times.

**Extended Data Figure 5.**
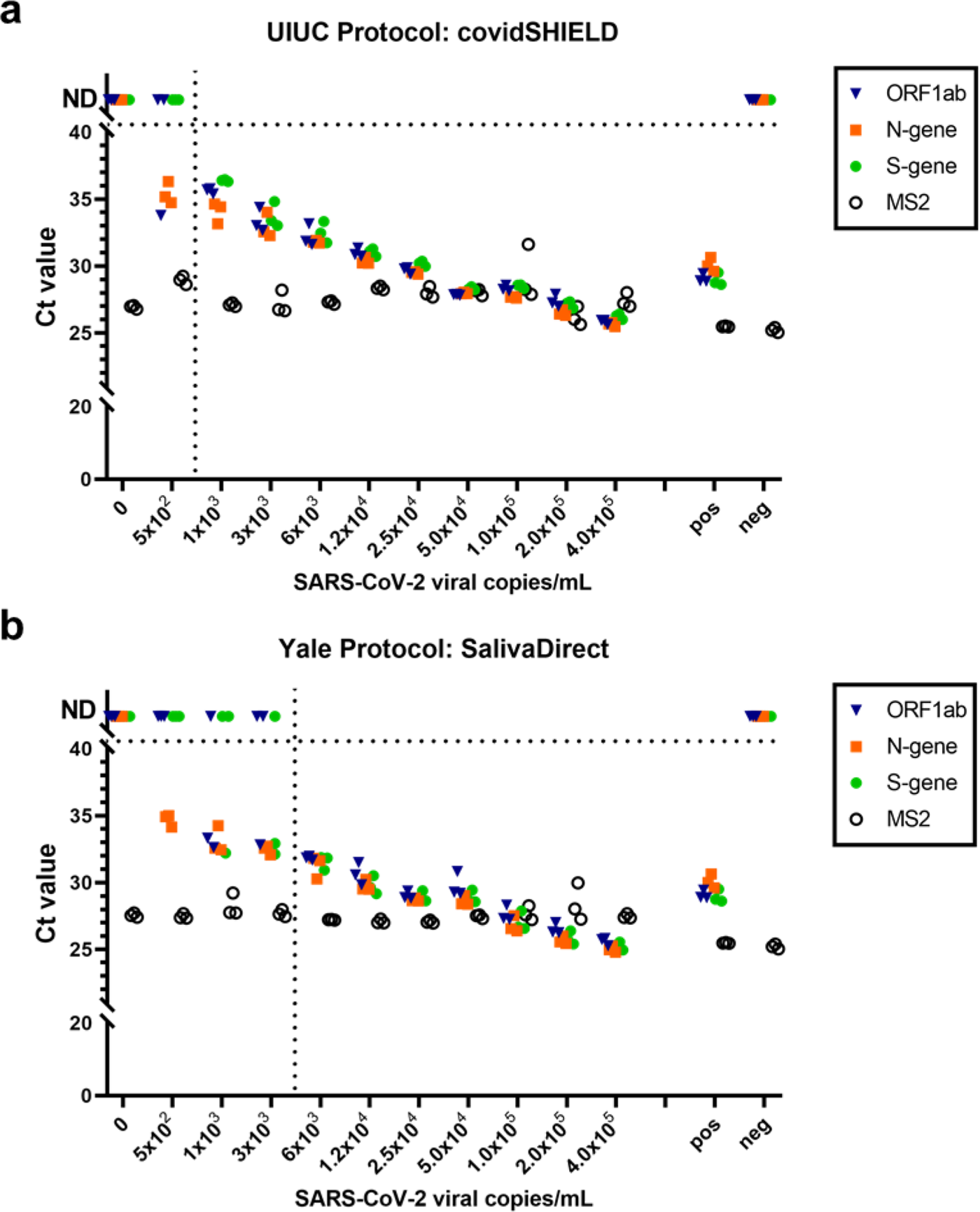
Comparison of covidSHIELD processing method (**a**, heat at 95°C for 30 minutes followed by TBE/tween addition) to SalivaDirect protocol (**b**,proteinase K treatment followed by heating at 95°C for 5 minutes). γ-irradiated SARS-CoV-2 was spiked into fresh human saliva (SARS-CoV-2 negative). All samples were analyzed with the ThermoFisher TaqPath COVID-19 combo kit on a QS7 instrument in triplicates together with a positive control (pos; SARS-CoV-2 positive control, 5.0×10^3^ copies/mL) and a negative control (neg; water). MS2 bacteriophage was added to the PCR reaction mix as internal control. This experiment was performed twice.

**Extended Data Figure 6.**
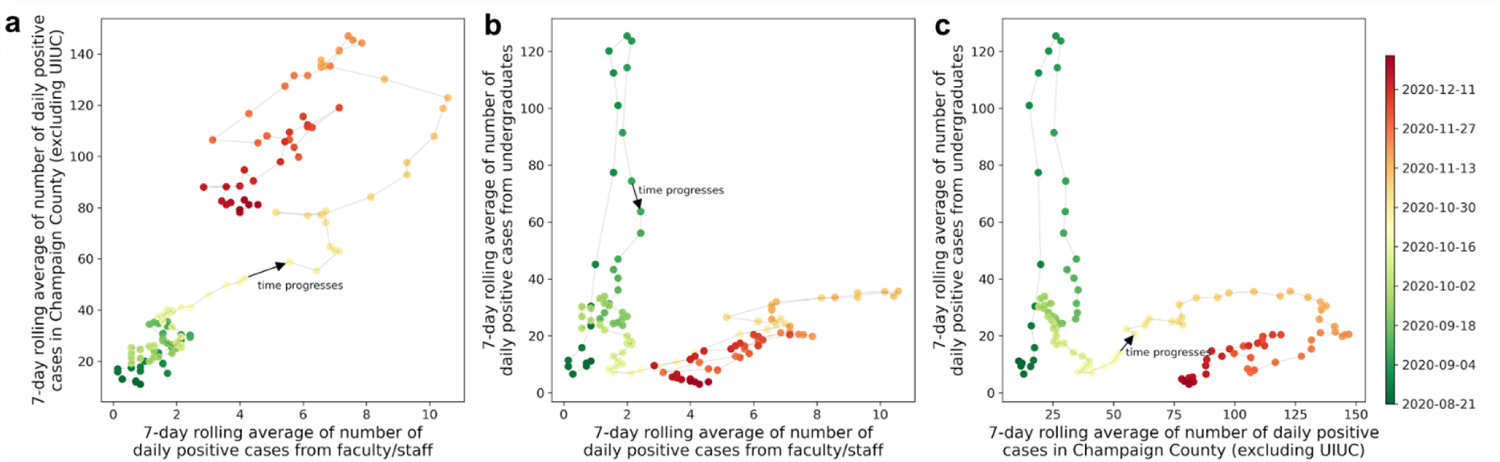
Measured correlations of the number of 7-day averaged daily new cases between residents in Champaign County, faculty/staff, and undergraduate students for the period between August 15 and December 23. All points in three scatter plots are colored according to their dates, as shown in the color bar on the right. (**a**) The number of 7-day averaged daily new cases of residents in Champaign county strongly correlated with that of faculty/staff especially for the period after October 18 (Pearson correlation coefficient 0.86, p-value 7.99×10^-38^). (**b**) At the beginning of the semester (around August 31) when there was a spike in daily new cases from undergraduate students, it had little influence on the faculty/staff. While at the late period of the semester (after October 18) when the case positivity in Illinois increased, they showed a correlation (Pearson correlation coefficient 0.88, p-value 1.33×10^-21^). (**c**) The initial spike in the number of daily new cases of undergraduate students doesn’t correlate with that of residents in Champaign county. While at the late period of the semester (after October 18), as the number of positive cases in Champaign county increased and then decreased, the number of cases of undergraduate students also showed a similar trend.

**Extended Data Figure 7.**
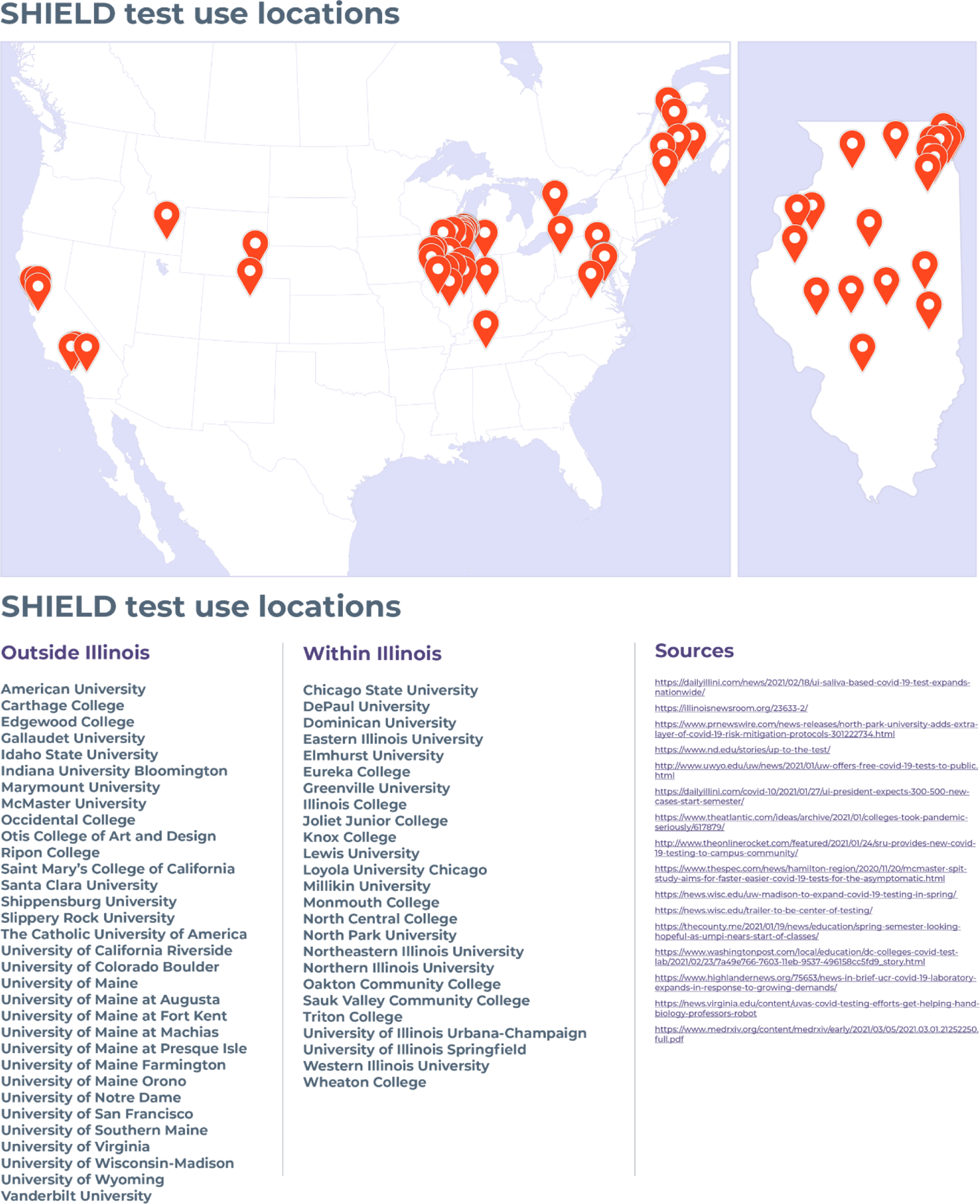
Some of the other colleges and universities now using the covidSHIELD saliva processing method to protect their communities.

**Extended Data Table 1.** Comparison of Ct values from 25 clinical samples that were split into two aliquots upon receipt, one set was processed using our covidSHIELD assay and the other set was subjected to RNA extraction using MagMax Viral/Pathogen II (MVP II) Nucleic Acid Isolation Kit (ThermoFisher).

| Sample # | Direct saliva Call | RNA extract Call | Direct saliva Ct Values |  |  |  | RNA extract Ct Values |  |  |  |
| --- | --- | --- | --- | --- | --- | --- | --- | --- | --- | --- |
|  |  |  | ORF1AB | N-gene | S-gene | MS2 | ORF1AB | N-gene | S-gene | MS2 |
| 1 | POSITIVE | POSITIVE | 34.87689 | 34.7379 | 35.15069 | 31.43486 | 32.53683 | 33.62223 | 33.32546 | 29.20287 |
| 2 | POSITIVE | POSITIVE | 35.77484 | 31.63383 | 34.52456 | 27.39116 | 31.82251 | 31.47362 | 33.69433 | 24.40539 |
| 3 | NEGATIVE | NEGATIVE | Undetermined | Undetermined | Undetermined | 31.98769 | Undetermined | Undetermined | Undetermined | 25.35025 |
| 4 | POSITIVE | POSITIVE | 21.35963 | 19.91812 | 21.34096 | Undetermined | 21.15125 | 20.97386 | 20.84411 | 25.25592 |
| 5 | POSITIVE | POSITIVE | 29.7138 | 28.12146 | 29.12307 | 36.42744 | 28.12993 | 28.94287 | 26.80732 | 23.59188 |
| 6 | POSITIVE | POSITIVE | 31.75227 | 31.18219 | 32.32317 | 30.34586 | 29.79028 | 30.34225 | 29.37346 | 23.5 |
| 7 | POSITIVE | POSITIVE | 27.67761 | 26.04979 | 27.44747 | 33.67909 | 24.12692 | 24.64138 | 23.57869 | 22.80227 |
| 8 | POSITIVE | POSITIVE | 36.4927 | 36.48437 | 36.93896 | 28.00187 | Undetermined | 34.4855 | 36.89221 | 22.77695 |
| 9 | POSITIVE | POSITIVE | 18.32149 | 16.63957 | 17.65472 | Undetermined | 14.88425 | 14.77061 | 15.18406 | 26.68707 |
| 10 | POSITIVE | POSITIVE | 30.50895 | 30.42821 | 31.71363 | 35.54698 | 28.93207 | 28.85836 | 29.24729 | 23.41111 |
| 11 | POSITIVE | POSITIVE | 32.64179 | 34.5071 | 36.60606 | 38.88396 | 32.00633 | 32.99638 | 29.73417 | 22.02418 |
| 12 | NEGATIVE | NEGATIVE | Undetermined | Undetermined | Undetermined | 30.05719 | Undetermined | Undetermined | Undetermined | 26.15957 |
| 13 | NEGATIVE | NEGATIVE | Undetermined | Undetermined | Undetermined | 30.69856 | Undetermined | Undetermined | Undetermined | 25.32277 |
| 14 | POSITIVE | POSITIVE | 24.31471 | 23.53031 | 24.30666 | 29.79528 | 23.33266 | 23.90056 | 23.92258 | 27.70757 |
| 15 | POSITIVE | POSITIVE | 18.86967 | 17.92603 | 19.15773 | Undetermined | 19.11883 | 19.35747 | 19.49679 | 25.0189 |
| 16 | POSITIVE | POSITIVE | Undetermined | 36.27135 | 34.65929 | 30.30633 | Undetermined | 34.78008 | 33.06602 | 24.60403 |
| 17 | POSITIVE | POSITIVE | 26.1404 | 23.63066 | 25.59911 | 31.9001 | 22.4836 | 22.44151 | 22.57868 | 23.68353 |
| 18 | POSITIVE | POSITIVE | 24.88006 | 24.41358 | 25.35321 | Undetermined | 24.94342 | 25.0757 | 25.15791 | 25.10076 |
| 19 | POSITIVE | POSITIVE | 34.05909 | 32.84715 | 33.15377 | 29.26073 | 30.61996 | 32.79143 | 30.9592 | 23.24416 |
| 20 | NEGATIVE | NEGATIVE | 31.63252 | 31.42906 | 33.89475 | 32.67826 | 34.07472 | 33.28623 | 35.13592 | 26.34162 |
| 21 | POSITIVE | POSITIVE | Undetermined | Undetermined | Undetermined | 29.93763 | Undetermined | Undetermined | Undetermined | 28.28117 |
| 22 | POSITIVE | POSITIVE | 33.39564 | 31.59387 | 33.96499 | 28.2531 | 28.93732 | 29.36552 | 29.37632 | 25.81484 |
| 23 | POSITIVE | POSITIVE | 17.52907 | 15.91184 | 17.7005 | Undetermined | 14.79202 | 14.88865 | 14.93889 | 31.47698 |
| 24 | POSITIVE | POSITIVE | 32.60616 | 29.36388 | 32.66383 | 30.71958 | 30.41657 | 31.16113 | 30.37413 | 26.24533 |
| 25 | POSITIVE | POSITIVE | 34.66038 | 32.72861 | 34.80508 | 30.83342 | 30.00526 | 31.25149 | 30.34891 | 24.93401 |
| **Average Ct for viral target genes** |  |  | **28.78** |  |  |  | **27.14** |  |  |  |

**Extended Data Table 2.**
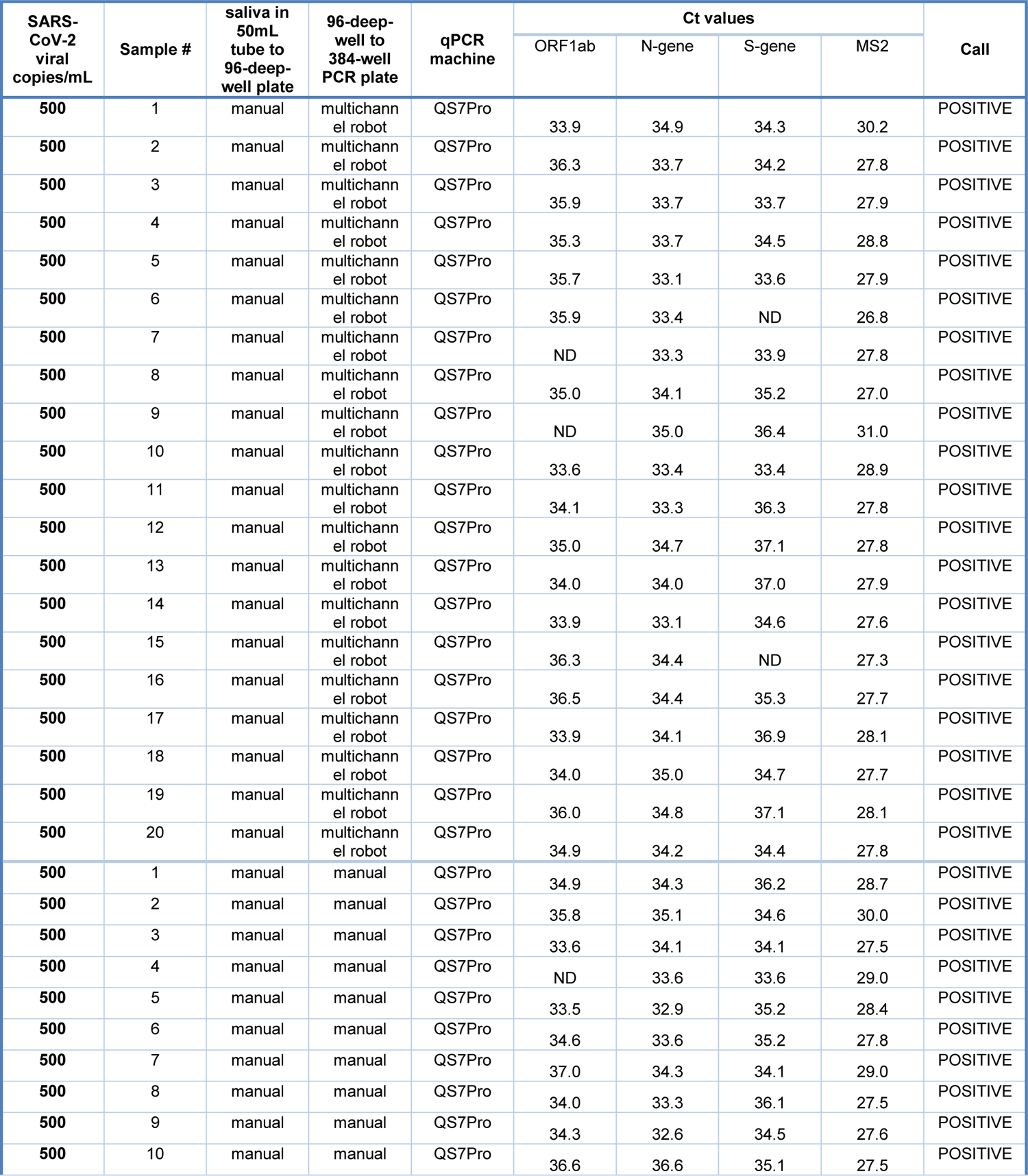
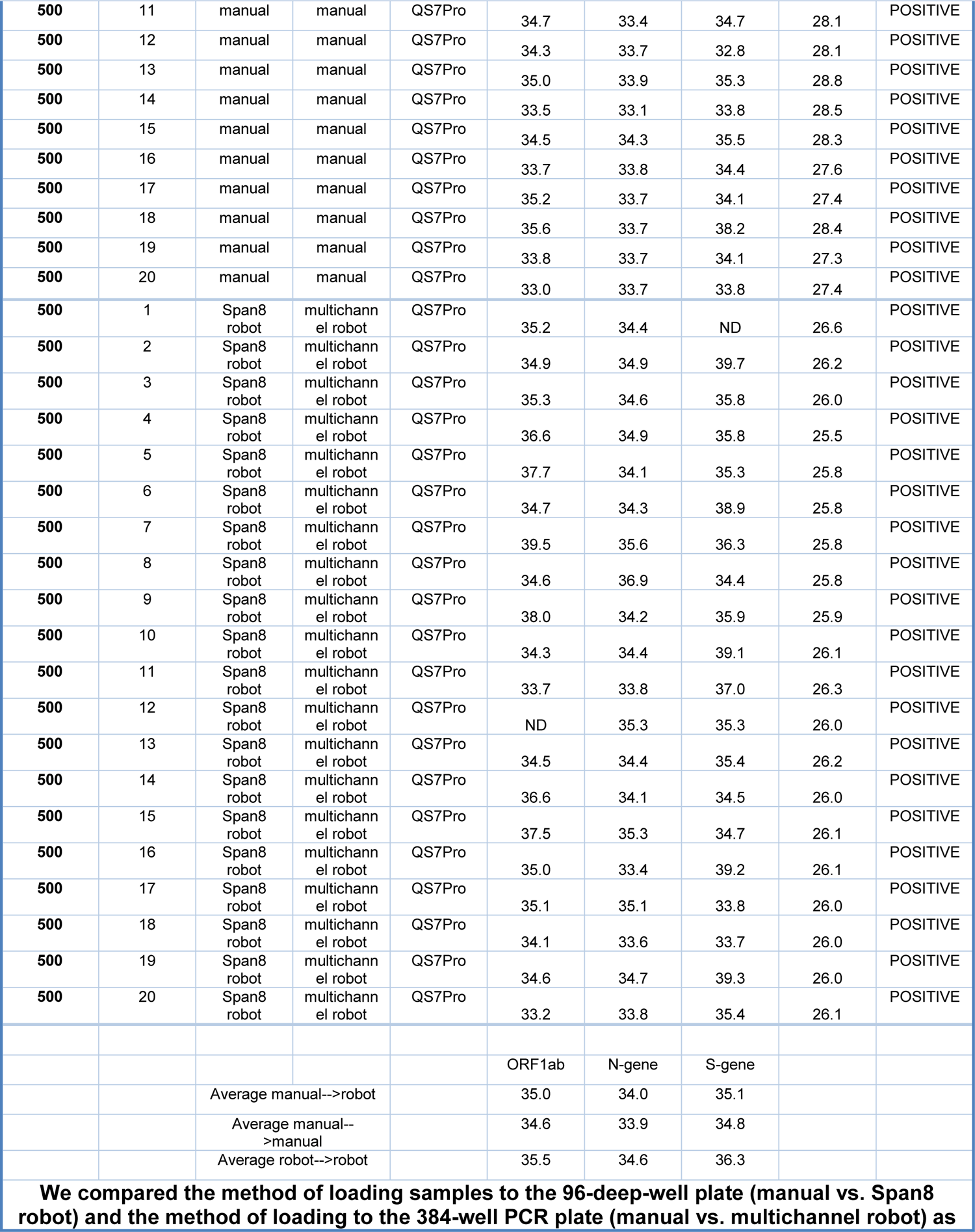
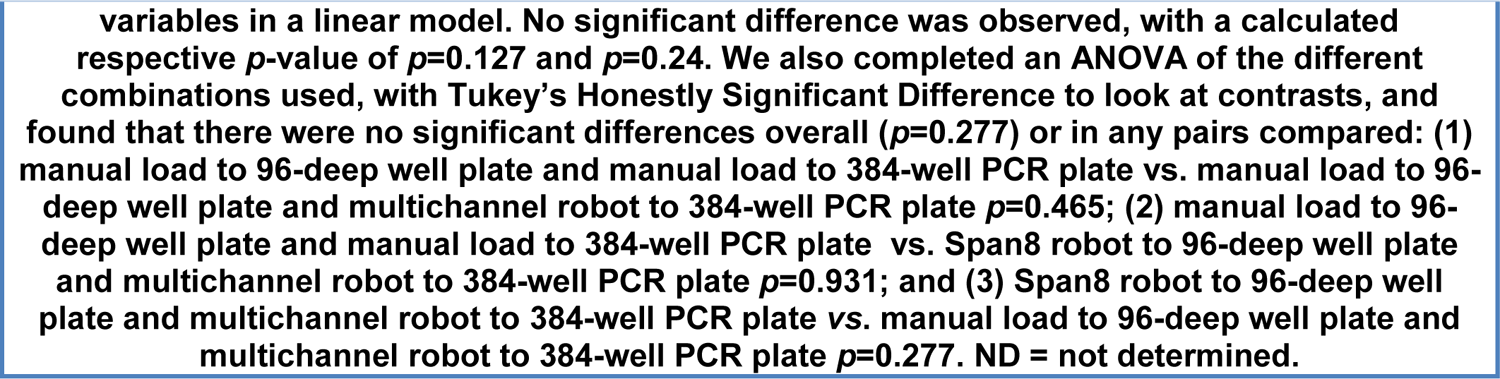
Comparison of method of loading to the 384-well PCR plate (manual vs. multichannel robot) and the method of loading heat-inactivated saliva samples to the 96-deep-well plates pre-loaded with 2xTBE/1% Tween-20 buffer (manual vs. Span8 robot).

**Extended Data Table 3.** Endogenous & Exogenous Interference testing results for covidSHIELD assay

| **Potential Interfering Substances** | **Concentration** | **Negative Samples** | **Positive Samples** |
| --- | --- | --- | --- |
|  |  | Detected | Detected |
| **Saliva Samples** |  |  |  |
| **Nasal congestion spray** | 15% v/v | 3/3 Negative | 3/3 Positive |
| **NeilMed Nasogel** | 1.25% | 3/3 Negative | 3/3 Positive |
| **Cepacol Lozenges (benzocaine/menthol)** | 3 mg/mL | 3/3 Negative | 3/3 Positive |
| **Chloroseptic Sore Throat spray** | 5% v/v | 3/3 Negative | 3/3 Positive |
| **Crest/Listerine Mouthwash** | 5% v/v | 3/3 Negative | 3/3 Positive |
| **Act dry mouth lozenges** | 3 mg/mL | 3/3 Negative | 3/3 Positive |
| **Toothpaste (Colgate)** | 0.5% v/v | 3/3 Negative | 1/3 Negative<br>2/3 Positive |
| **Mucin: bovine submaxillary gland, type I-S** | 2.5 mg/ml | 3/3 Negative | 3/3 Positive |
| **Human Genomic DNA** | 10 ng/μl | 3/3 Negative | 3/3 Positive |
| **White blood cells/Leukocytes** | 1 to 5x10<sup>6</sup> cells/mL | 3/3 Negative | 3/3 Positive |
| **Nicotine** | 0.03 mg/mL | 3/3 Negative | 3/3 Positive |

**Extended Data Table 4.**
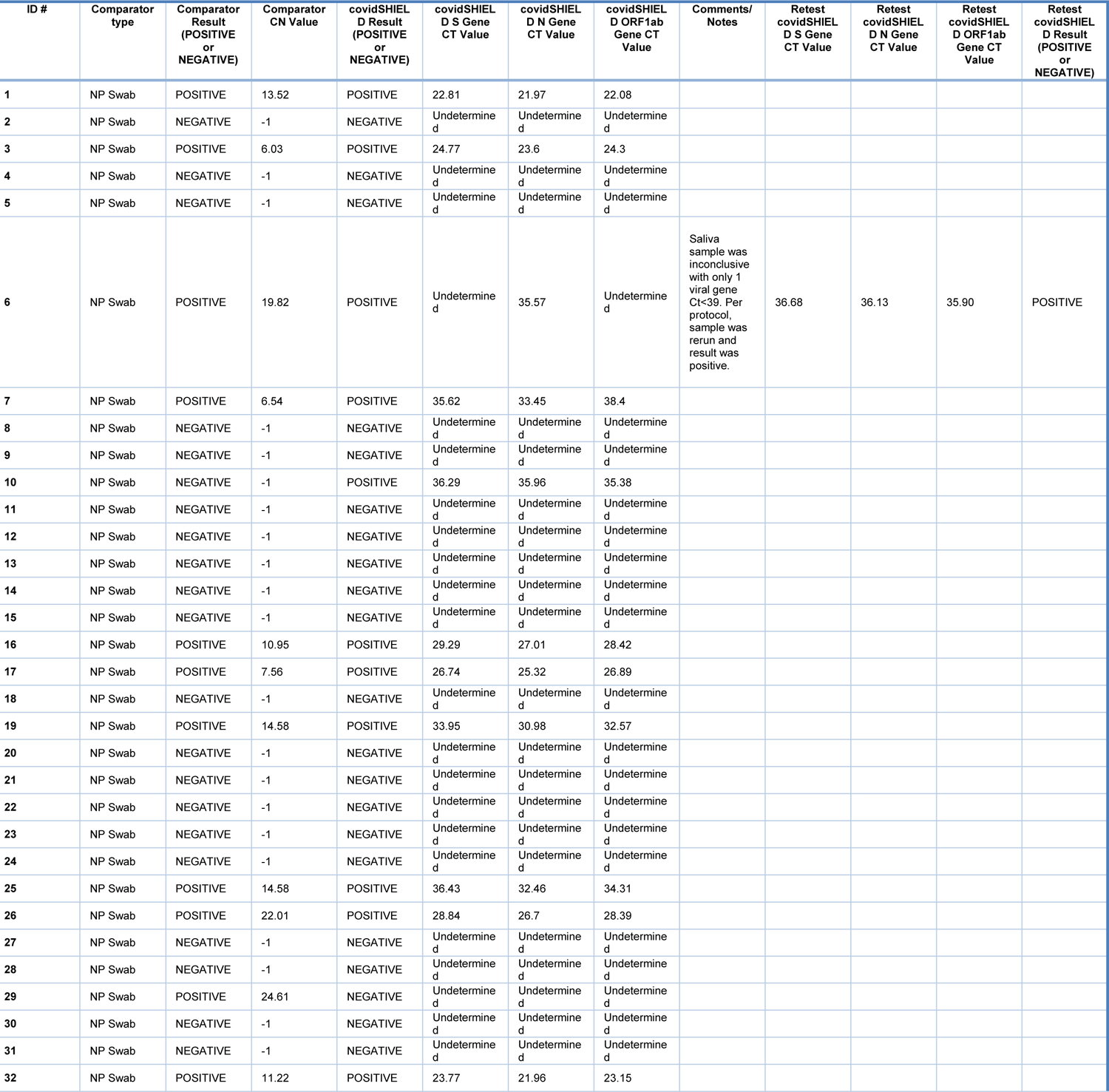
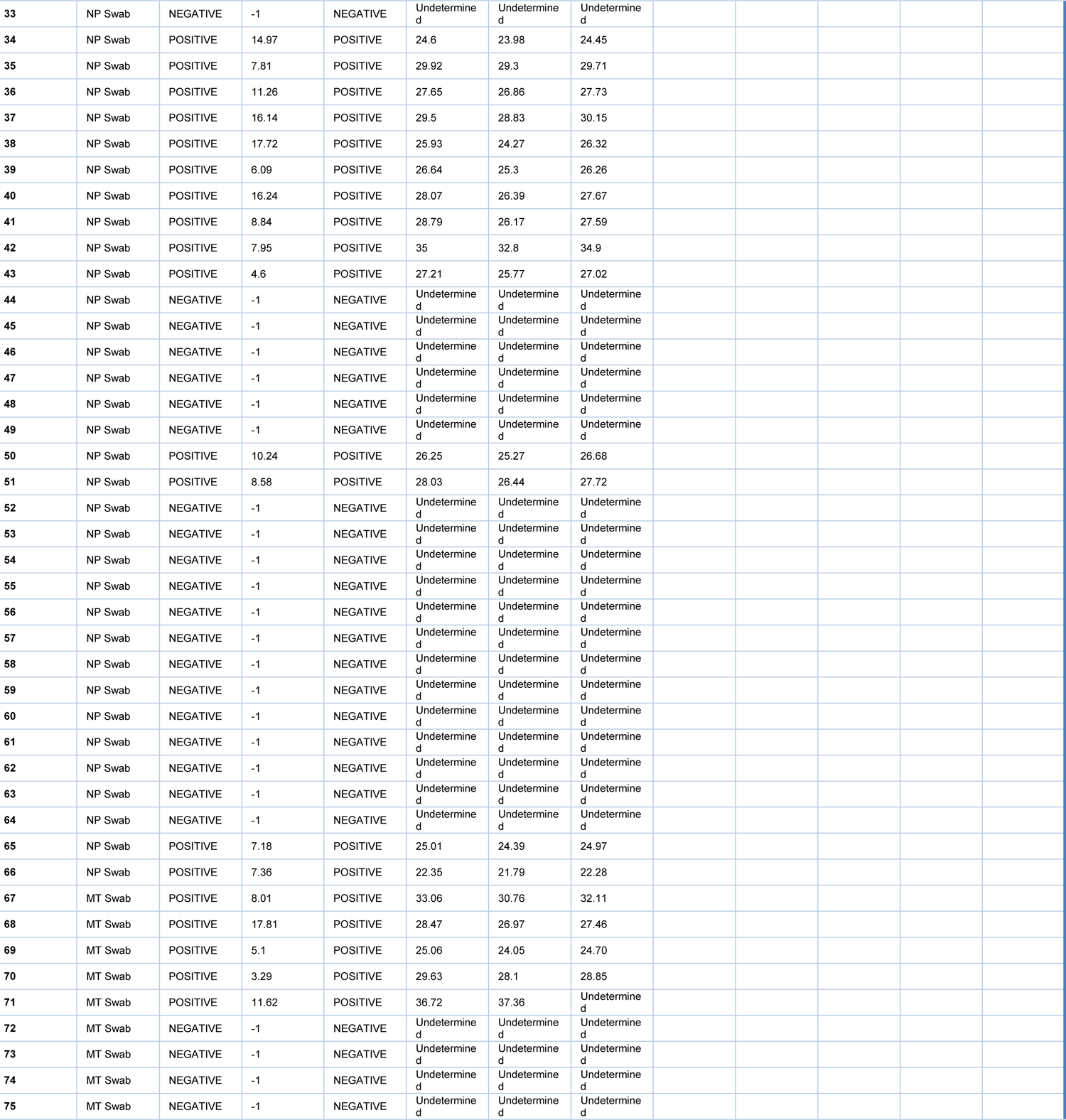
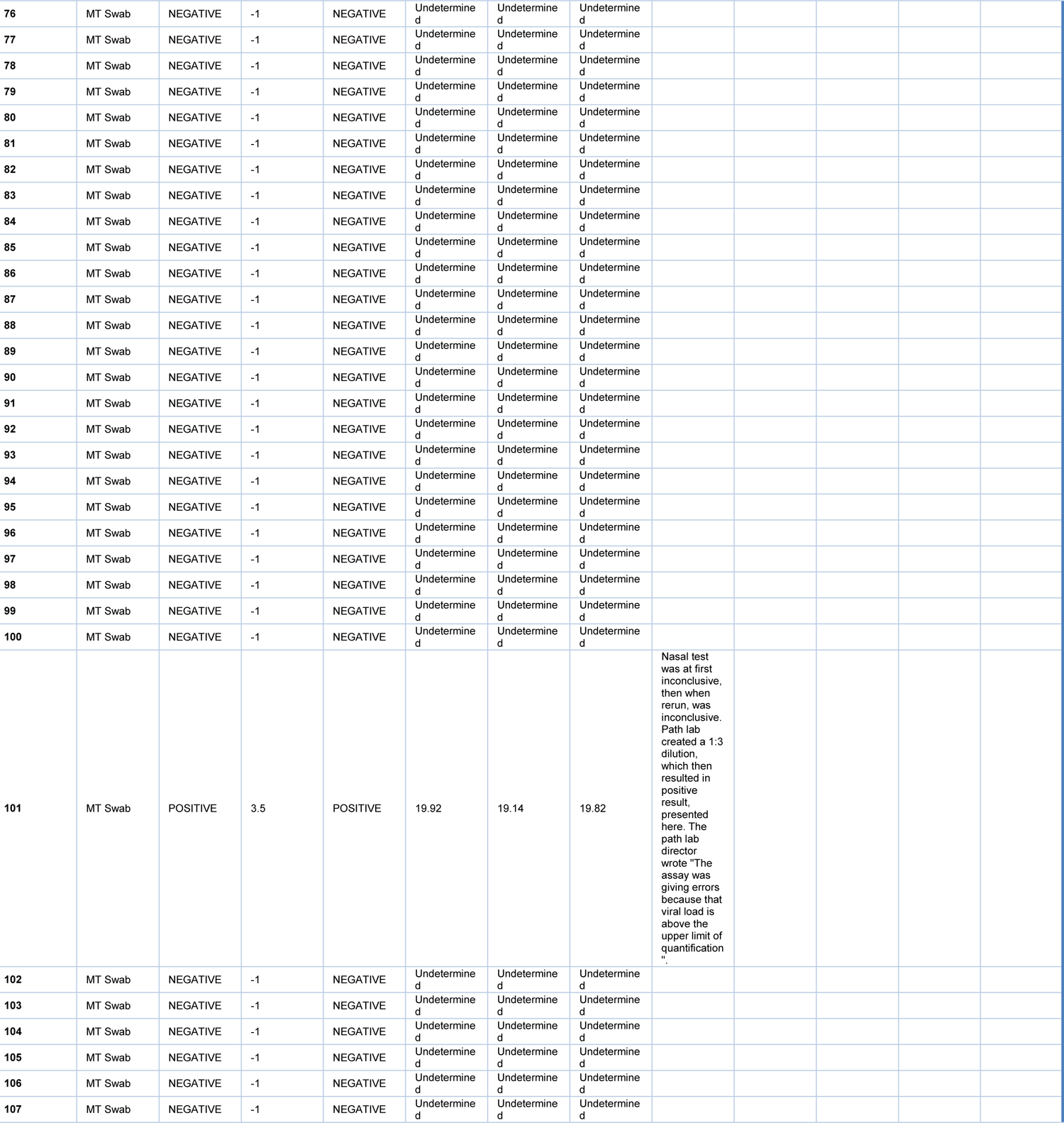
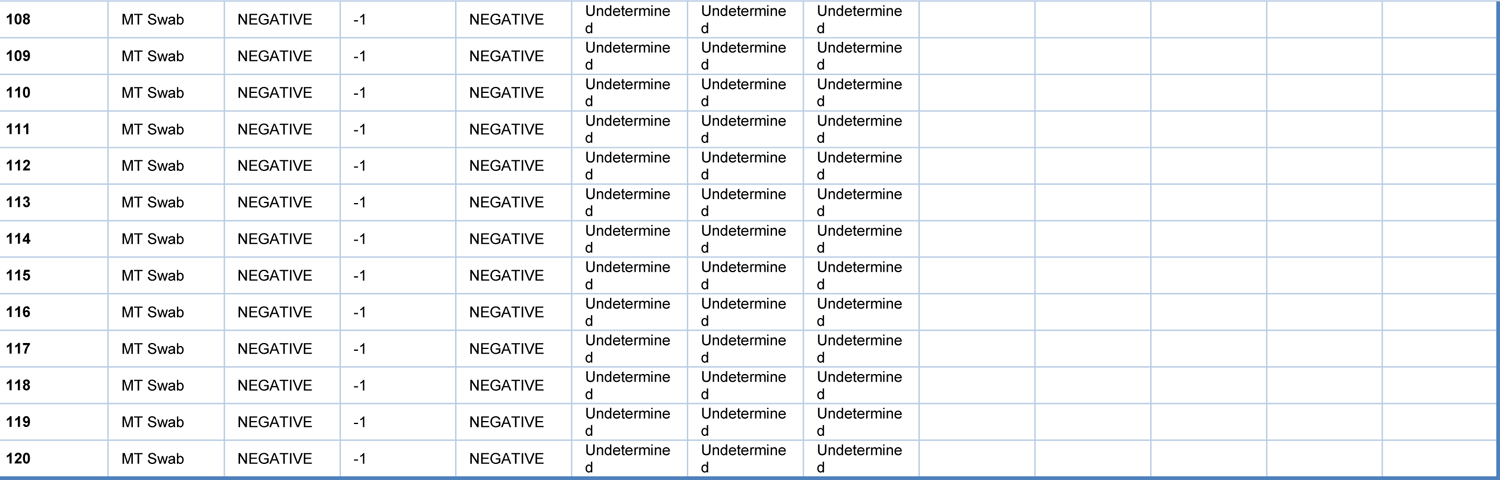
Qualitative outcome of parallel testing of paired mid-turbinate swabs and saliva with the Abbott RealTime SARS-CoV-2 assay and covidSHIELD.

**Extended Data Table 5.** Method comparison study completed to support the correlation between saliva samples processed with covidSHIELD and nasal samples processed with Abbott RealTime SARS-CoV-2 assay performed on the Abbott m2000 System.

|  |  |
| --- | --- |
| **Name of Candidate device** | **covidSHIELD** |
| **Study Title** | Method comparison study for covidSHIELD to an RT-qPCR nasopharyngeal or mid-turbinate reference method for detection of SARS-CoV-2. |
| **Study Type (CLIA – high/moderate complexity, CLIA waived/POC, OTC, usability, other)** | The candidate device is a high complexity assay only housed in high complexity CLIA-certified laboratories<br>Saliva samples were self-collected with observation |
| **Number of Study Sites** | Samples were collected at 4 different sites (one in Urbana, IL; One in Madison, WI; two in Chicago, IL); candidate testing was done at one testing site. |
| **Operators** | Operators processed samples for comparator and candidate in high complexity labs with appropriate trained personnel. |
| **Target Population** | Study population included:<br>Participants over the age of 18<br>Participants suspected of COVID-19 (either with symptoms or with known exposures to someone positive for COVID-19). |
| **Number of Participants** | We enrolled a total of 120 participants. We had 31 positives and 89 negatives as determined by the comparator (either an MT or NP swab, as available at each site). |
| **Study Objective** | To determine the positive percent agreement (PPA) and negative percent agreement (NPA) between the candidate device on saliva samples and an EUA-authorized RT-qPCR assay (specifically, the Abbott RealTime SARS-CoV-2 assay with an LoD of 2,700 NDU/ml according to the list of FDA Reference Panel assays). |
| **Study Comparator** | Abbott RealTime SARS-CoV-2 assay performed on the Abbott m2000 System |
| **Comparator Device Sample matrix and collection method** | The comparator sample was either a mid-turbinate or nasopharyngeal swab collected by a healthcare professional. Each swab was placed into a vial of collection media and transported to the test instrument (per the comparator manufacturer's instructions). |
| **Study General Design and Duration** | Participants were approached:<br>• Prospectively while visiting a saliva collection site (Urbana) OR<br>• Prospectively while visiting one of two different urgent care centers (Chicago) OR<br>• Following a recent anterior nares swab positive test for SARS-CoV-2. Approximately 24 hours following the anterior nares swab collection, those individuals who tested positive were asked to consent to a research study. (Madison)<br>After consent, participants answered survey questions about symptoms and provided a self-collected saliva specimen and either a mid-turbinate or nasopharyngeal swab specimen (as available at each site), collected by a healthcare professional. |
| **Participant & Sample Inclusion / Exclusion Criteria** | **Inclusion Criteria for Participants:**<br>Participants meeting the following criteria were included in this study:<br>1. Participant is 18 years of age or older<br>2. Participant must be willing and able to provide a saliva sample and willing and able to have a healthcare professional successfully collect a mid-turbinate or nasopharyngeal sample<br>3. Participant must be willing and able to answer questions about their symptoms as part of the data collected for this study<br>4. Participant self-reported being symptomatic<br>**Exclusion Criteria for Participants:**<br>Patients with the following criteria were excluded:<br>1. Participants under 18 years of age<br>2. Participants who have previously tested positive for SARS-CoV-2, indicating that they have previously been infected with SARS-CoV-2. |
|  | 3. Participants who have had anything “by mouth” (eaten, drank, brushed their teeth, smoked or chewed gum etc.) in the last 60 minutes.<br>**Exclusion Criteria for Samples:**<br>1. If sample tube label was illegible, the sample would have been excluded from the study<br>2. If sample tube had evidence of leaking or volume loss, the sample would have been excluded from the study<br>3. If sample had visual debris present or any obvious contamination due to blood, food or beverage etc, the sample would have been excluded from the study. |
| **Study Procedures** | • Participants were instructed to provide a saliva sample by allowing saliva to collect in the mouth and gently expelling saliva into the collection tube. Sample donors then capped their tube. The participant handed the sample to the healthcare professional or collection site staff who placed the tube in a collection container.<br>• The participant was asked survey questions about their symptoms.<br>• The healthcare professional collected the mid-turbinate or nasopharyngeal sample and transferred the swab into the collection media (per the comparator manufacturer’s instructions).<br>• Both samples were transported (via ground) to the testing location. All saliva samples were tested at the UIUC CLIA-certified lab on the candidate test; all swabs were transported by ground to the University of Illinois Chicago Hospital where they were tested with the Abbott RealTime SARS-CoV-2 assay performed on the Abbott m2000 System. |
| **Statistical Analysis** | • Positive percent agreement (PPA) and negative percent agreement (NPA) were determined by comparing results from the candidate test with those from the comparator test. |

**Extended Data Table 6.** Details to Capture on Case Report Form

|  |  |
| --- | --- |
| **Testing of External Controls** | Candidate Assay:<br>For every test run performed, a known positive and negative control was included in the covidSHEILD test's reaction plate, as well as internal controls for each sample. The negative control consisted of UltraPure DNase- and RNase-free water, in order to evaluate cross-contamination between loading samples into the 384-well reaction plate. The positive control consisted of the Thermo Fisher TaqPath COVID-19 Combo Kit provided positive control, in order to evaluate the RT-qPCR reagents (primers, probes, Master Mix) and the ability of the QuantStudio 4, 7 Flex, and 7 Pro systems to detect viral genes. The internal control consisted of the Thermo Fisher TaqPath COVID-19 Combo Kit provided MS2, which was spiked into the MasterMix reaction, in order to evaluate if material within the saliva interfered with the detection of viral RNA by RT-qPCR. Controls were included on every plate. Beckman Biomek i5 Span8 and Multichannel robotics were used to transfer samples for some of the tests; for other tests, samples were transferred via manual pipetting.<br>The test results were evaluated according to the interpretation tables in the EUA Summary (https://www.fda.gov/media/146317/download).<br>Comparator Assay:<br>The control protocol was followed according to standard practice in the clinical laboratory and per manufacturer's instructions. |
| **Sample Storage for candidate device** | Saliva samples were stored and/or shipped at room temperature for no longer than 7 days in the capped sample collection tube prior to testing. |

**Extended Data Table 7.** Comparison of mid-turbinate (MT) swab and saliva from 17 individuals identified with low viral load based on MT swab analyzed using Abbott Alinity RT-PCR.

| Sample # | Mid-turbinate swab |  | Saliva |  |  |  |
| --- | --- | --- | --- | --- | --- | --- |
|  | Alinity Ct | Result | S-gene Ct | N-gene Ct | ORF1ab-gene Ct | Result |
| 1 | 32.83 | positive | 34.07 | 33.39 | 35.19 | positive |
| 2 | 36.41 | positive | 33.14 | 31.68 | 32.72 | positive |
| 3 | 35.91 | positive | 29.49 | 28.94 | 29.7 | positive |
| 4 | 40.13 | positive | 34.61 | 32.19 | 34.71 | positive |
| 5 | 39.97 | positive | 30.52 | 29.82 | 30.59 | positive |
| 6 | 38.63 | positive | 35.09 | 33.68 | 35.27 | positive |
| 7 | 37.45 | positive | 34.66 | 33.12 | 33.94 | positive |
| 8 | 37.04 | positive | 33.87 | 32.2 | 33.26 | positive |
| 9 | 35.66 | positive | 27.19 | 25.79 | 27.38 | positive |
| 10 | 37.68 | positive | 35.17 | 34.77 | 37.61 | positive |
| 11 | 37.74 | positive | 34.99 | 33.81 | 34.65 | positive |
| 12 | 40.47 | positive | 33.93 | 33.99 | 36.53 | positive |
| 13 | 33.36 | positive | undetermined | 36.26 | undetermined | inconclusive |
| 14 | 32.48 | positive | 21.75 | 21.34 | 21.37 | positive |
| 15 | 32.48 | positive | 28.17 | 27.72 | 28.23 | positive |
| 16 | 41.73 | positive | 35.95 | 34.23 | undetermined | positive |
| 17 | 39.83 | positive | 37.41 | 39.66 | 36.55 | positive |

**Extended Data Table 8.** Quanta emission rates, and ventilation rates in different zone types. How many zones in each type is given in the “Count” column

| **Zone type** | **Count** | **$E_i$**<br>**(*quanta/hour*)** | **$r_{vent}$**<br>**(*hour<sup>-1</sup>*)** |
| --- | --- | --- | --- |
| Bar | 20 | 150 | 15 |
| Restaurant | 200 | 20 | 10 |
| Cafe | 50 | 15 | 10 |
| Library | 50 | 4 | 3.5 |
| Classroom | 790 | Professor<br>:100 | 3.5 |
|  |  | Student:4 |  |
| Dorm | 300 | 4 | 3.5 |
| Dorm parties | 300 | 150 | 3.5 |

**Extended Data Table 9:**
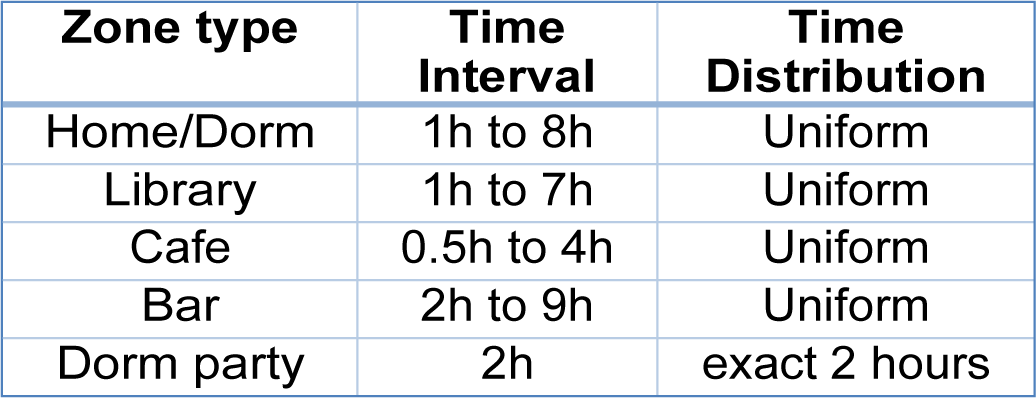
Assumption for how long agents stay in different zones

| **Zone type** | **Time Interval** | **Time Distribution** |
| --- | --- | --- |
| Home/Dorm | 1h to 8h | Uniform |
| Library | 1h to 7h | Uniform |
| Cafe | 0.5h to 4h | Uniform |
| Bar | 2h to 9h | Uniform |
| Dorm party | 2h | exact 2 hours |

## Notes

### Clinical Trial

Western IRB-approved protocol number 20203538

### Author Declarations

Western IRB-approved protocol number 20203538

## References

1. Richmond, C. S., Sabin, A. P., Jobe, D. A., Lovrich, S. D. & Kenny, P. A. SARS-CoV-2 sequencing reveals rapid transmission from college student clusters resulting in morbidity and deaths in vulnerable populations. medRxiv, 2020.2010.2012.20210294, doi:10.1101/2020.10.12.20210294 (2020).

2. Wilson, E. et al. Multiple COVID-19 Clusters on a University Campus — North Carolina, August 2020. MMWR. Morbidity and Mortality Weekly Report 69, 1416-1418, doi:10.15585/mmwr.mm6939e3 (2020).

3. Johansson, M. A. et al. SARS-CoV-2 Transmission From People Without COVID-19 Symptoms. JAMA Netw Open 4, e2035057, doi:10.1001/jamanetworkopen.2020.35057 (2021).

4. Ye, F. et al. Delivery of infection from asymptomatic carriers of COVID-19 in a familial cluster. Int. J. Infect. Dis. 94, 133–138, doi:10.1016/j.ijid.2020.03.042 (2020).

5. Shental, N. et al. Efficient high throughput SARS-CoV-2 testing to detect asymptomatic carriers. medRxiv, doi:10.1101/2020.04.14.20064618 (2020).

6. Furukawa, N. W., Brooks, J. T. & Sobel, J. Evidence Supporting Transmission of Severe Acute Respiratory Syndrome Coronavirus 2 While Presymptomatic or Asymptomatic. Emerg. Infect. Dis. 26, doi:10.3201/eid2607.201595 (2020).

7. Li, R. et al. Substantial undocumented infection facilitates the rapid dissemination of novel coronavirus (SARS-CoV-2). *Science (New York*, N.Y.) 368, 489–493, doi:10.1126/science.abb3221 (2020).

8. He, X. et al. Temporal dynamics in viral shedding and transmissibility of COVID-19. Nat. Med. 26, 672–675 (2020).

9. Larremore, D. B. et al. Test sensitivity is secondary to frequency and turnaround time for COVID-19 surveillance. medRxiv, doi:10.1101/2020.06.22.20136309 (2020).

10. Wölfel, R. et al. Virological assessment of hospitalized patients with COVID-2019. Nature 581, 465–469, doi:10.1038/s41586-020-2196-x (2020).

11. Tan, W. et al. Viral kinetics and antibody responses in patients with COVID-19. MedRxiv (2020).

12. Zhou, R. et al. Viral dynamics in asymptomatic patients with COVID-19. Int. J. Infect. Dis. 96, 288–290, doi:10.1016/j.ijid.2020.05.030 (2020).

13. Goyal, A., Reeves, D. B., Cardozo-Ojeda, E. F., Schiffer, J. T. & Mayer, B. T. Viral load and contact heterogeneity predict SARS-CoV-2 transmission and super-spreading events. Elife 10, doi:10.7554/eLife.63537 (2021).

14. Ferretti, L. et al. Quantifying SARS-CoV-2 transmission suggests epidemic control with digital contact tracing. *Science (New York*, N.Y.) 368, doi:10.1126/science.abb6936 (2020).

15. Han, P. & Ivanovski, S. Saliva—Friend and Foe in the COVID-19 Outbreak. Diagnostics 10, 290, doi:10.3390/diagnostics10050290 (2020).

16. Sharma, S., Kumar, V., Chawla, A. & Logani, A. Rapid detection of SARS-CoV-2 in saliva: can an endodontist take the lead in point-of-care COVID-19 testing? Int. Endod. J. 53, 1017–1019, doi:10.1111/iej.13317 (2020).

17. Braz-Silva, P. H., Pallos, D., Giannecchini, S. & To, K. K. w. SARS-CoV-2: What can saliva tell us? Oral Diseases, doi:10.1111/odi.13365 (2020).

18. Wyllie, A. L. et al. Saliva is more sensitive for SARS-CoV-2 detection in COVID-19 patients than nasopharyngeal swabs. *Medrxiv* (2020).

19. Iwasaki, S. et al. Comparison of SARS-CoV-2 detection in nasopharyngeal swab and saliva. J. Infect. 81, e145–e147, doi:10.1016/j.jinf.2020.05.071 (2020).

20. Czumbel, L. M. et al. Saliva as a Candidate for COVID-19 Diagnostic Testing: A Meta-Analysis. Front Med (Lausanne) 7, 465, doi:10.3389/fmed.2020.00465 (2020).

21. Pasomsub, E. et al. Saliva sample as a non-invasive specimen for the diagnosis of coronavirus disease 2019: a cross-sectional study. Clinical Microbiology and Infection 27, 285.e281–285.e284, doi:10.1016/j.cmi.2020.05.001 (2021).

22. Butler-Laporte, G. et al. Comparison of Saliva and Nasopharyngeal Swab Nucleic Acid Amplification Testing for Detection of SARS-CoV-2: A Systematic Review and Meta-analysis. JAMA internal medicine 181, 353–360, doi:10.1001/jamainternmed.2020.8876 (2021).

23. Ranoa, D. R. E. et al. Saliva-Based Molecular Testing for SARS-CoV-2 that Bypasses RNA Extraction. bioRxiv, 2020.2006.2018.159434, doi:10.1101/2020.06.18.159434 (2020).

24. Batéjat, C., Grassin, Q., Manuguerra, J.-C. & Leclercq, I. Heat inactivation of the Severe Acute Respiratory Syndrome Coronavirus 2. Cold Spring Harbor Laboratory, doi:10.1101/2020.05.01.067769 (2020).

25. Fomsgaard, A. S. & Rosenstierne, M. W. An alternative workflow for molecular detection of SARS-CoV-2 - escape from the NA extraction kit-shortage. medRxiv, 2020.2003.2027.20044495, doi:10.1101/2020.03.27.20044495 (2020).

26. Vogels, C. B. F. et al. SalivaDirect: A simplified and flexible platform to enhance SARS-CoV-2 testing capacity. Med 2, 263–280.e266, doi:https://doi.org/10.1016/j.medj.2020.12.010 (2021).

27. Bullard, J. et al. Predicting infectious SARS-CoV-2 from diagnostic samples. Clin. Infect. Dis., doi:10.1093/cid/ciaa638 (2020).

28. Vetter, P. et al. Daily Viral Kinetics and Innate and Adaptive Immune Response Assessment in COVID-19: a Case Series. mSphere 5, doi:10.1128/mSphere.00827-20 (2020).

29. Perera, R. A. et al. SARS-CoV-2 virus culture from the upper respiratory tract: Correlation with viral load, subgenomic viral RNA and duration of illness. medRxiv, 2020.2007.2008.20148783, doi:10.1101/2020.07.08.20148783 (2020).

30. Cevik, M. et al. SARS-CoV-2, SARS-CoV, and MERS-CoV viral load dynamics, duration of viral shedding, and infectiousness: a systematic review and meta-analysis. Lancet Microbe 2, e13–e22, doi:10.1016/S2666-5247(20)30172-5 (2021).

31. Gniazdowski, V. et al. Repeat COVID-19 Molecular Testing: Correlation of SARS-CoV-2 Culture with Molecular Assays and Cycle Thresholds. Clin. Infect. Dis., doi:10.1093/cid/ciaa1616 (2020).

32. Wallinga, J. & Lipsitch, M. How generation intervals shape the relationship between growth rates and reproductive numbers. Proc. Biol. Sci. 274, 599–604, doi:10.1098/rspb.2006.3754 (2007).

33. Cori, A., Ferguson, N. M., Fraser, C. & Cauchemez, S. A new framework and software to estimate time-varying reproduction numbers during epidemics. Am. J. Epidemiol. 178, 1505–1512, doi:10.1093/aje/kwt133 (2013).

34. Centers for Disease, C. & Prevention, C.-R. (2021).

35. The American Community Survey, T. P. (2019).

36. CDC SVI Documentation 2018. (2020).

37. Buonanno, G., Stabile, L. & Morawska, L. Estimation of airborne viral emission: Quanta emission rate of SARS-CoV-2 for infection risk assessment. Environment International 141, 105794, doi:https://doi.org/10.1016/j.envint.2020.105794 (2020).

38. Marr, L., et al. FAQs on Protecting Yourself from aerosol transmission. (2020).

39. Stetzenbach, L. D., Buttner, M. P. & Cruz, P. Detection and enumeration of airborne biocontaminants. Current opinion in biotechnology 15, 170–174, doi:10.1016/j.copbio.2004.04.009 (2004).

40. Elbanna, A. et al. Entry screening and multi-layer mitigation of COVID-19 cases for a safe university reopening. Epidemiology, doi:10.1101/2020.08.29.20184473 (2020).

41. safer-illinois-app v. 2.0 (2020).

42. Rokwire. (2020).

43. Safer Illinois. (2020).

